# Severe cognitive decline in long-term care is related to gut microbiome production of metabolites involved in neurotransmission, immunomodulation, and autophagy

**DOI:** 10.1101/2023.03.06.23286878

**Authors:** Andrew P. Shoubridge, Lucy Carpenter, Erin Flynn, Lito E. Papanicolas, Josephine Collins, David Gordon, David J. Lynn, Craig Whitehead, Lex E.X. Leong, Monica Cations, David P. De Souza, Vinod K. Narayana, Jocelyn M. Choo, Steve L. Wesselingh, Maria Crotty, Maria C. Inacio, Kerry Ivey, Steven L. Taylor, Geraint B. Rogers

## Abstract

Ageing-associated cognitive decline affects more than half of those in long-term residential aged care. Emerging evidence suggests that gut microbiome-host interactions influence the effects of modifiable risk factors. We investigated the relationship between gut microbiome characteristics and severity of cognitive impairment (CI) in 159 residents of long-term aged care. Severe CI was associated with a significantly increased abundance of proinflammatory bacterial species, including *Methanobrevibacter smithii* and *Alistipes finegoldii*, and decreased relative abundance of beneficial bacterial clades. Severe CI was associated with increased microbial capacity for methanogenesis, and reduced capacity for synthesis of short-chain fatty acids, neurotransmitters glutamate and gamma-aminobutyric acid, and amino acids required for neuro-protective lysosomal activity. These relationships were independent of age, sex, antibiotic exposure, and diet. Our findings implicate multiple gut microbiome-brain pathways in ageing-associated cognitive decline, including inflammation, neurotransmission, and autophagy, and highlight the potential to predict and prevent cognitive decline through microbiome-targeted strategies.

## Introduction

Progressive loss of cognitive function is a common feature of ageing and is not limited to those with dementia (Albert et al., 1995; Comijs et al., 2004; Harada et al., 2013; Verdi et al., 2018). Contributory pathologies, often occurring in combination, include ischaemic or haemorrhagic infarcts within the brain (characteristic of vascular dementia) (Henon et al., 2001; Ye et al., 2015), the accumulation of amyloid plaques and neurofibrillary tangles (characteristic of Alzheimer’s disease) (Bussian et al., 2018; Luo et al., 2020), and the development of abnormal collections of alpha-synuclein protein within diseased brain neurons (characteristic of Lewy body dementia) (Lin et al., 2017; Mattila et al., 2000). While these pathophysiological processes are increasingly well-characterised, the factors that contribute to them and their relationship to external risk exposures remain poorly understood.

In addition to genetic factors (Koistinaho et al., 2004; Lin et al., 2018; Liu et al., 2017; Parcon et al., 2018; Shi et al., 2017; Ulrich et al., 2018), modifiable risk factors associated with dementia have been identified. Modifiable risk factors include exposures (smoking, excessive alcohol consumption, physical inactivity, air pollution, diet); health conditions (hypertension, obesity, depression, diabetes, traumatic brain injury, hearing impairment), and social factors (less education, and low social contact) (Livingston et al., 2020). Together, these modifiable risk factors are estimated to account for 40% of dementia incidence (Livingston et al., 2020). Identifying how such factors influence the pathophysiology of ageing-associated cognitive impairment (CI) is essential to the development of effective prevention and treatment.

The gut microbiome influences neurophysiology, central nervous system, and cognitive function through discrete bidirectional pathways, collectively termed the microbiome-gut-brain axis (Cryan et al., 2019; Rogers et al., 2016; Sharon et al., 2016; Shoubridge et al., 2022). These pathways include the microbial synthesis of neurotransmitters, such as gamma-aminobutyric acid (GABA), noradrenaline, dopamine and serotonin (Valles-Colomer et al., 2019; Yano et al., 2015), the modulation of systemic immunity (Correa-Oliveira et al., 2016; Dalile et al., 2019), and metabolism of essential amino acids, such as tyramine and tryptophan (Lai et al., 2021; Marx et al., 2021). They also involve production of immune and metabolically active metabolites, such as short-chain fatty acids (SCFAs) and 4-ethylphenylsulfate, and activation of nerve growth factor, glial-derived neurotrophic factor, and brain-derived neurotrophic factor secretion (Bonfili et al., 2021; Soto et al., 2018). Such microbial traits have the potential to contribute substantially to the development of neurological diseases, including Alzheimer’s (Kim et al., 2020; Vogt et al., 2017), Huntington’s (Bjorkqvist et al., 2008; Du et al., 2020; Wasser et al., 2020), and Parkinson’s diseases (Sampson et al., 2016; Sun et al., 2018).

Ageing-associated gut microbiome characteristics (Claesson et al., 2011; Meyer et al., 2022) are linked to progressive frailty and cognitive decline (Komanduri et al., 2021; Manderino et al., 2017; Meyer et al., 2022; Verdi et al., 2018). External exposures that disrupt the microbiome, such as antibiotics, can further contribute to altered neurological homeostasis and poorer cognitive outcomes (Desbonnet et al., 2015; Frohlich et al., 2016; Lynn et al., 2021). In contrast, dietary interventions that alter the composition of the gut microbiome in a beneficial manner can result in improvements in cognitive function (Ghosh et al., 2020). Such findings suggest that the relationship between the gut microbiome and host neurophysiology may provide a basis to predict and/or prevent the onset and progression of ageing-associated cognitive decline. Potential causality in these relationships is suggested by studies that have successfully recapitulated impairment of memory and synaptic plasticity following faecal microbiota transplant from aged mice to younger mice (D’Amato et al., 2020).

Our aim was to explore whether the severity of CI experienced by residents of long-term aged care facilities (sometimes referred to as nursing homes, care homes, or residential aged care facilities) is associated with characteristics of the gut microbiome, and if so, whether such relationships might provide mechanistic insight into CI pathogenesis.

## Materials and Methods

### Study design, cohort and data collection

The Generating evidence on Resistant bacteria in the Aged Care Environment (GRACE) study (www.gracestudy.com.au) was a cohort study supported by the Australian Medical Research Future Fund (Grant No. GNT1152268). Ethical approval for the study was obtained from the Southern Adelaide Clinical Human Research Ethics Committee (HREC/18/SAC/244). The GRACE study investigated the carriage and transfer of resistant bacteria in long-term aged care facilities and was conducted between 2018 and 2020. GRACE enrolled 279 residents in five long-term aged care facilities in metropolitan South Australia. Anonymised participant data, including assessments of cognition and behaviour, were collected via an entry into care funding assessment (Aged Care Funding Instrument [ACFI]), in addition to medications prescribed via the Pharmaceutical Benefits Scheme (PBS) (Carpenter, 2021).

### Assessment of cognitive impairment

The *Cognitive Skills* component of the ACFI was used as a basis for assessment of cognitive impairment. This cognitive skills component assesses a person’s cognitive abilities in everyday activities, including memory, self-care, and orientation (AIHW, 2002; Department of Health and Ageing, 2016), as defined via the Psychogeriatric Assessment Scales – Cognitive Impairment Scales (PAS-CIS) method (Jorm et al., 1995). Where individuals were unable to undertake the PAS-CIS test, for example, non-English speaking, sensory impairment, or severe cognitive impairment beyond the scope of the instrument, the ACFI cognitive skills assessment was based on a clinical report by a registered health professional (Department of Health and Ageing, 2016). The ACFI cognitive skills component utilised the PAS-CIS and/or clinical reports to rate an individual’s level of cognitive impairment as none or minimal (PAS-CIS = 0-3), mild (4-9), moderate (10-15), or severe (16-21).

### Cognitive impairment cohort

GRACE participants were categorised according to their cognitive skills rating, as defined in the ACFI. Participants were excluded if: 1) their cognitive skills assessment was not completed or missing, 2) the date of stool collection was not known, 3) the date of the cognitive assessment was not known, 4) the participant was diagnosed with a developmental or intellectual disability, or 5) the period between cognitive skills assessment and stool sample collection was not known or was deemed an outlier (>1462 days as determined using the ROUT method of regression and outlier removal (Motulsky & Brown, 2006)). A total of 45 participants were excluded (detailed in **Supplementary** Fig. 1). Participants with dementia and a missing PAS-CIS rating were imputed the median PAS-CIS value from their cognitive skills assessment group. Mental and behavioural diagnoses of dementia, depression, and delirium were ascertained from the ACFI, where a documented diagnosis from a medical practitioner was provided.

### Faecal collection, DNA extraction, metagenomic sequencing and bioinformatics

Stool was collected and stored using Norgen Stool Nucleic Acid Collection and Preservation Tubes (Norgen Biotek, ON, Canada) and microbial DNA extracted using the PowerLyzer PowerSoil DNA Isolation Kit (Qiagen, Hilden, Germany) as described previously (Carpenter, 2021). Indexed, paired-end DNA libraries were prepared using the Nextera XT DNA Library Prep Kit (Illumina, CA, USA), as per manufacturer’s instructions. Samples were sequenced at a depth of 5 Gb on an Illumina Novaseq platform with 150bp paired-end reads. Forward and reverse sequences were quality-filtered using Trimmomatic (v0.39) and human reads were removed with Bowtie (v2.3.5.1) against the NCBI human reference genome release GRCh38 (Bolger et al., 2014; Langmead & Salzberg, 2012). Taxonomic relative abundance was assigned using MetaPhlAn (v3.0) (Beghini et al., 2021), while microbial metabolic pathway abundance was assigned using HUMAnN (v3.0) against the MetaCyc database (Beghini et al., 2021). Sequence data has been entered into the European Nucleotide Archive (ENA) at EMBL-EBI under accession number PRJEB51408.

### Microbiome characterisation

The taxonomic relative abundance at the species level was used to generate alpha diversity (within-group) and beta diversity (between-group) measures. Alpha diversity measures included Pielou’s evenness (J’: a score between 0-1 where scores are influenced more by the evenness of abundant species), the Shannon-Wiener diversity (H’): a score of the number and equal representation of different types of species (Peet, 1974), and species richness (d: total number of unique species identified per participant), and were generated using the ‘vegan’ R package (Oksanen, 2022).

The Bray-Curtis index was calculated to compare microbiome similarity between groups (beta diversity), using square-root transformed species relative abundance data (PRIMER 6 (v6.1.16)). For sensitivity analysis, weighted and unweighted UniFrac distance matrices (Lozupone & Knight, 2005) were calculated using the “calculate_unifrac” MetaPhlAn R script (Beghini et al., 2021). Non-metric multidimensional scaling (nMDS) plots for all beta diversity measures were generated using the ‘vegan’ package in R and visualised using ‘ggplot2’.

### Microbial functional profiling

The functional capacity of the gut microbiota was defined by the genetically encoded functional traits detected within the metagenome. These MetaCyc pathways from HUMAnN were filtered to only analyse those present in >30% of participants. Two functional profiling analyses were performed: an untargeted analysis of all filtered pathways (n=400), and a targeted analysis based on pathways with a hypothesised functional role in cognitive impairment (n=70). These included pathways involved in neurotransmitter biosynthesis (n=2), SCFA production (n=25), and amino acid biosynthesis (n=43).

### Metabolite profiling

As a confirmatory analysis of microbial functional capacity, the metabolomic profile of a randomly selected subgroup of individuals (n=35; n=11-12/group) was established. Stool metabolite analysis was performed on an Agilent 1200 series high-performance liquid chromatography system (Agilent Technologies) (methods modified from (Gubert et al., 2022; Kong et al., 2021) and detailed in **Supplementary Text 1**). Briefly, metabolite extraction and analysis were performed separately for SCFAs and polar metabolites. SCFAs analysis was performed using an Agilent 6490 series triple quadrupole mass spectrometer (Agilent Technologies) while polar metabolites (a screen for 165 low molecular weight metabolites, e.g. amino acids) were analysed using an Agilent 6545 series quadrupole time-of-flight mass spectrometer (Agilent Technologies). Resultant data matrices were imported to the web-based platform MetaboAnalyst (v5.0) for quality control checks. SCFA data were normalised to internal standards, and polar metabolite data were log-transformed and median-normalised.

### Covariates

Covariates were: days since cognitive assessment (below or equal/above the median), age (low, medium or high tertile), sex (male or female), medication history (Pharmaceutical Benefits Scheme (PBS) data available or unavailable), medications that affect gastrointestinal health and are prevalent in aged care facilities: antibiotic use (yes or no); proton pump inhibitor use (yes or no); opioid use (yes or no); laxative use (yes or no), meal texture at time of cognitive assessment (regular or soft/smooth), and liquid texture at time of cognitive assessment (normal/thin or thick). Medication use was defined as two or more supplies to a resident within 90 days prior to stool collection.

### Statistical analysis

Both unadjusted and multivariate regression models were applied in all analyses. Multivariate adjusted models accounted for time between cognitive assessment and stool collection, age, sex, antibiotics, proton pump inhibitors, opioids, laxatives, meal texture, and liquid texture (as detailed above).

Beta diversity analysis was performed using permutational multivariate analyses of variance (PERMANOVA) on Bray-Curtis, weighted, and unweighted UniFrac distance matrices in an unrestricted permutation of raw data. Only the Bray-Curtis metric was assessed with the multivariate-adjusted model. PERMANOVA analyses were performed using PRIMER 6, with 9999 permutations.

Within-individual microbiome variables included alpha diversity, phyla-level relative abundance (only those detected in >30% of participants), species-level relative abundance (only those detected in >30% of participants), metabolic pathway abundance (only those detected in >30% of participants), and metabolite intensities. All within-individual variables were converted to groups consisting of: zero values, tertile 1, tertile 2, and tertile 3. Ordinal logistic regression was performed to assess the effect of CI on microbiome variables using the ‘MASS’ function in R. The odds ratios and 95% confidence intervals for the coefficients of the regression models were calculated and tested for statistical significance (p<0.05) as CI severity increased, using the PAS-CIS score of cognitive impairment as the predictor variable. False Discovery Rate (FDR) multiple hypothesis testing was conducted with the Benjamini and Hochberg method across all profiles using the ‘p.adjust’ function in R, at a significant threshold of 0.05. Correlations between microbial functional capacity and detected metabolites were calculated by two-tailed Spearman correlations and tested for statistical significance (p<0.05).

## Results

The study group of 159 participants did not differ from the original GRACE cohort in any of the assessed characteristics (**Supplementary Table 1**). CI was classified as mild in 46 individuals (28.9%) with a median PAS-CIS score of 6.6 (IQR=5.0-8.0), moderate in 58 (36.5%; PAS-CIS median=11.0; IQR=11.0-12.8), and severe in 55 (34.6%; PAS-CIS median=18.0; IQR=17.0-18.8) (**Table 1; Supplementary** Fig. 2A). The median time from assessment of cognitive skills to the subsequent collection of stool samples was 437 days (IQR: 185-741). Participant age, sex ratio, time since cognitive assessment, and use of antibiotics, opioids, and laxatives, did not differ significantly between CI severity categories (p>0.05; **Supplementary** Fig. 2B-F). However, the number of days that an individual had been residing in long-term aged care was significantly higher for those in the severe CI group (median=939 days; IQR=219-854) compared to the mild CI group (median=500 days; IQR=130-627; p<0.05). Proton pump inhibitor usage was significantly lower in the severe CI group (p<0.05). Within the severe CI group, 53/55 (96.4%) had a concurrent diagnosis of dementia, 33/58 (56.9%) of moderate CI had a dementia diagnosis, and 4/46 (8.7%) of those with mild CI.

**Table 1.**
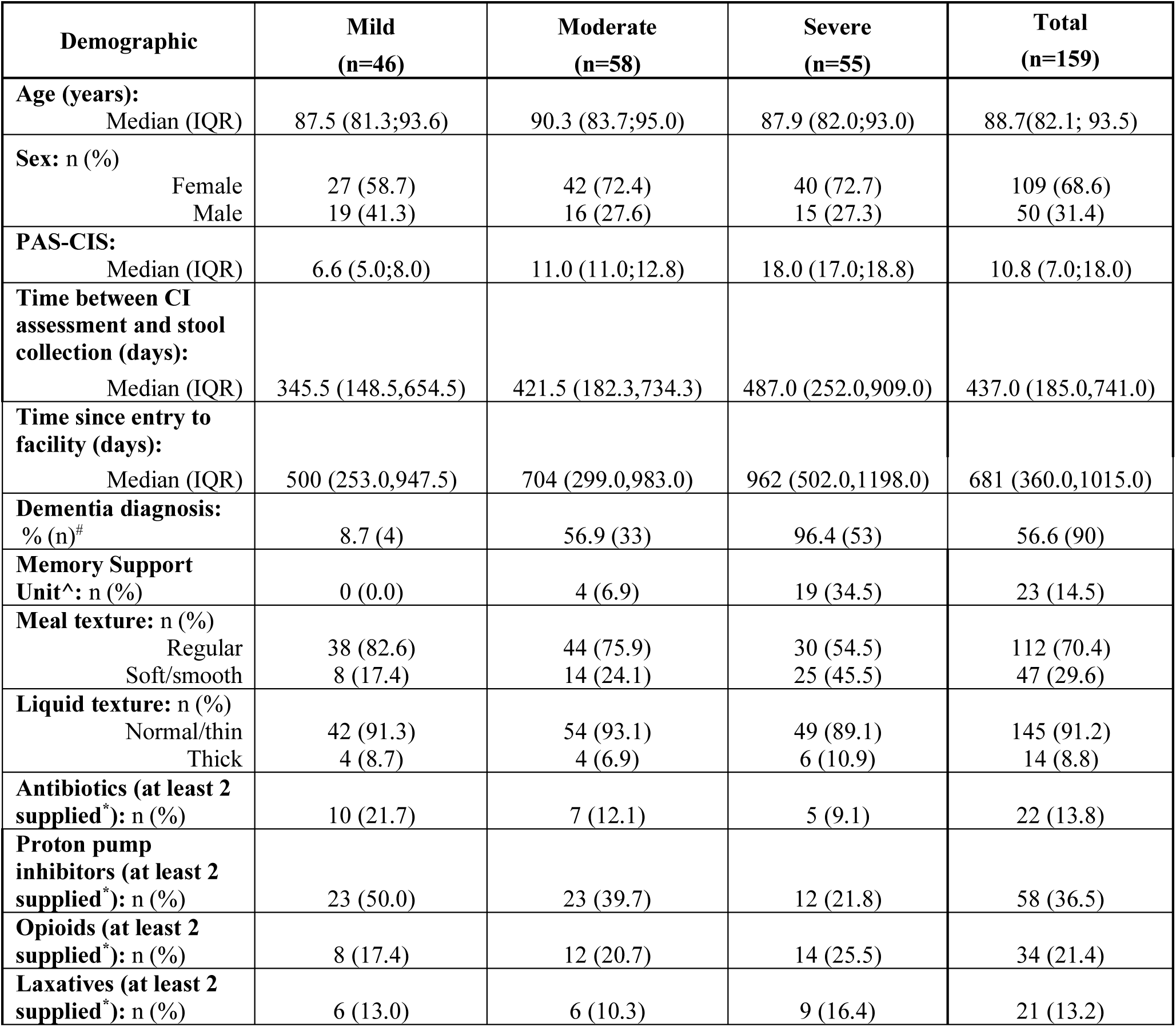
Study Cohort Characteristics by Severity of Cognitive Impairment. ^#^Extracted from Aged Care Funding Instrument data. ^Memory support units (also known as dementia units/wards, memory care, or special care units) are secure areas within long-term care facilities specially designed to accommodate residents with dementia. *Medication use defined as two or more supplies of the same medication within 90 days prior to stool collection.

### Gut microbiome characteristics differ by CI severity

Gut microbiome characterisation of long-term aged care residents with CI was determined by metagenomic sequencing of collected stool samples (**Fig. 1A**). A total of 11 bacterial phyla were detected across the 159 stool samples, consisting of 186 genera (586 species). The composition and distribution of taxa was broadly similar with previous studies of aged populations (Claesson et al., 2011; Jackson et al., 2016), with high representation of *Eggerthella lenta, Escherichia coli*, *Faecalibacterium prausnitzii*, and *Clostridium* species (**Fig. 1B**), and genera within the Bacteroidota (formerly Bacteroides) and Bacillota (Firmicutes) phyla (**Fig. 1C**).

**Figure 1.**
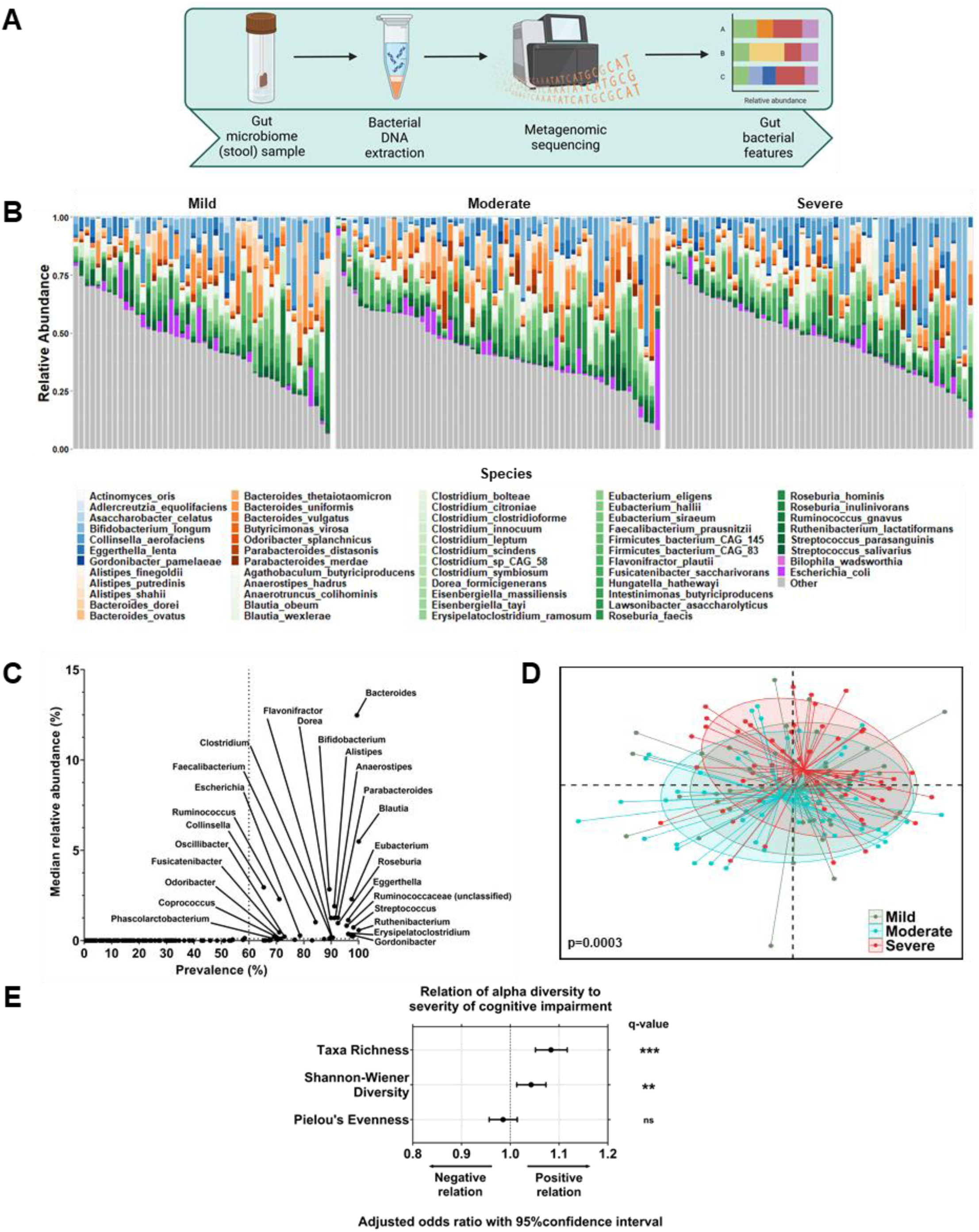
Gut microbiome of residents of long-term aged care facilities stratified by cognitive impairment (CI). **A**) Characterisation of the gut microbiome of long-term aged care residents with cognitive impairment determined by metagenomic sequencing of collected stool samples. **B**) Taxa bar plot of core species grouped by cognitive impairment severity (present in >60% of participants). Species coloured by phyla: Actinomycetota = blues; Bacteroidota = oranges; Bacillota = greens; Pseudomonadota = purples; non-core species (other) = grey. **C**) The frequency of genera detected and their median relative abundances, labelled with core genera. **D**) Non-metric multidimensional scaling plot of Bray-Curtis similarity matrix, grouped by CI severity (mild, n=46; moderate, n=58; severe, n=55), showing significant divergence between CI groups following multivariate analysis (p(perm)=0.0003). **E**) Odds ratio and 95% confidence interval of effect of CI severity on microbiome diversity (taxa richness, Shannon-Wiener diversity, and Pielou’s evenness), following multivariate analysis. Multivariate analysis adjusted for time since CI assessment, age, sex, antibiotic use, proton pump inhibitor use, opioid use, laxative use, recorded medical history, meal texture, and liquid texture. ns=not significant; **q<0.01; ***q<0.001 for adjusted p-values following FDR correction.

Following adjustment for time since cognitive assessment, age, sex, medication use, and diet, the faecal microbiota composition differed significantly between mild, moderate, and severe CI (p(perm)=0.0003; R^2^=2.21%; **Fig. 1D**; **Table 2**). This difference was greatest between severe CI and mild CI (p(perm)=0.0023), and severe CI and moderate CI (p(perm)=0.0003), and consistent with the unadjusted model (**Table 2**). Repeated analysis using weighted and unweighted UniFrac dissimilarity did not identify significant intergroup differences, apart from between mild and moderate CI groups using weighted UniFrac dissimilarity (unadjusted p(perm)=0.037; **Supplementary** Fig. 4).

**Table 2.** Permutational ANOVA of the gut microbiome by severity of cognitive impairment. ^a^Time since cognitive impairment assessment + age + sex+ antibiotic use + proton pump inhibitor use + opioid use + laxative use + recorded medication history + meal texture + liquid texture. ^Permutation p-value generated with a PERMANOVA. *Degrees of freedom = 2.

|  | Unadjusted Model |  |  | Multivariate-adjusted Model<sup>a</sup> |  |  |
| --- | --- | --- | --- | --- | --- | --- |
| Main Test | Pseudo-*F* Ratio | *R*<sup>2</sup> | p(perm)<sup>^</sup> | Pseudo-*F* Ratio | *R*<sup>2</sup> | p(perm)<sup>^</sup> |
| Cognitive Impairment* | 1.9646 | 0.0246 | **0.0002** | 1.815 | 0.0221 | **0.0003** |
| Pairwise Test | *t* |  | p(perm)<sup>^</sup> | *t* |  | p(perm)<sup>^</sup> |
| Mild v Moderate | 1.0596 |  | 0.2583 | 1.1135 |  | 0.1478 |
| Mild v Severe | 1.4474 |  | **0.0017** | 1.4228 |  | **0.0023** |
| Moderate v Severe | 1.6304 |  | **0.0002** | 1.5265 |  | **0.0003** |

Analysis of microbiota diversity identified a positive association between CI severity and taxa richness (odds ratio (OR): 1.08 [95% confidence interval 1.05, 1.12], q<0.001) and Shannon-Wiener diversity (OR: 1.043 [1.013, 1.073], q<0.05; **Fig. 1E**). However, there was no association between CI severity and Pielou’s evenness (OR: 0.985 [0.957, 1.014], q>0.05; **Fig. 1E**).

To assess whether specific taxa differed with CI, phylum-level and species-level relative abundances were assessed. Of the seven phyla present in at least 30% of participants, five differed significantly with CI (**Fig. 2**). Pseudomonadota (Proteobacteria) (OR: 0.937 [0.909, 0.965], q<0.001) and Bacillota (OR: 0.943 [0.915, 0.971], q<0.001) were lower with increasing CI severity (**Fig. 2**). In contrast, Euryarchaeota (OR: 1.097 [1.065, 1.131], q<0.001), Actinomycetota (Actinobacteria) (OR: 1.066 [1.035, 1.099], q<0.001), and Synergistota (Synergistetes) (OR: 1.043 [1.008, 1.079], q<0.05) were higher with increasing CI severity (**Fig. 2**).

**Figure 2.**
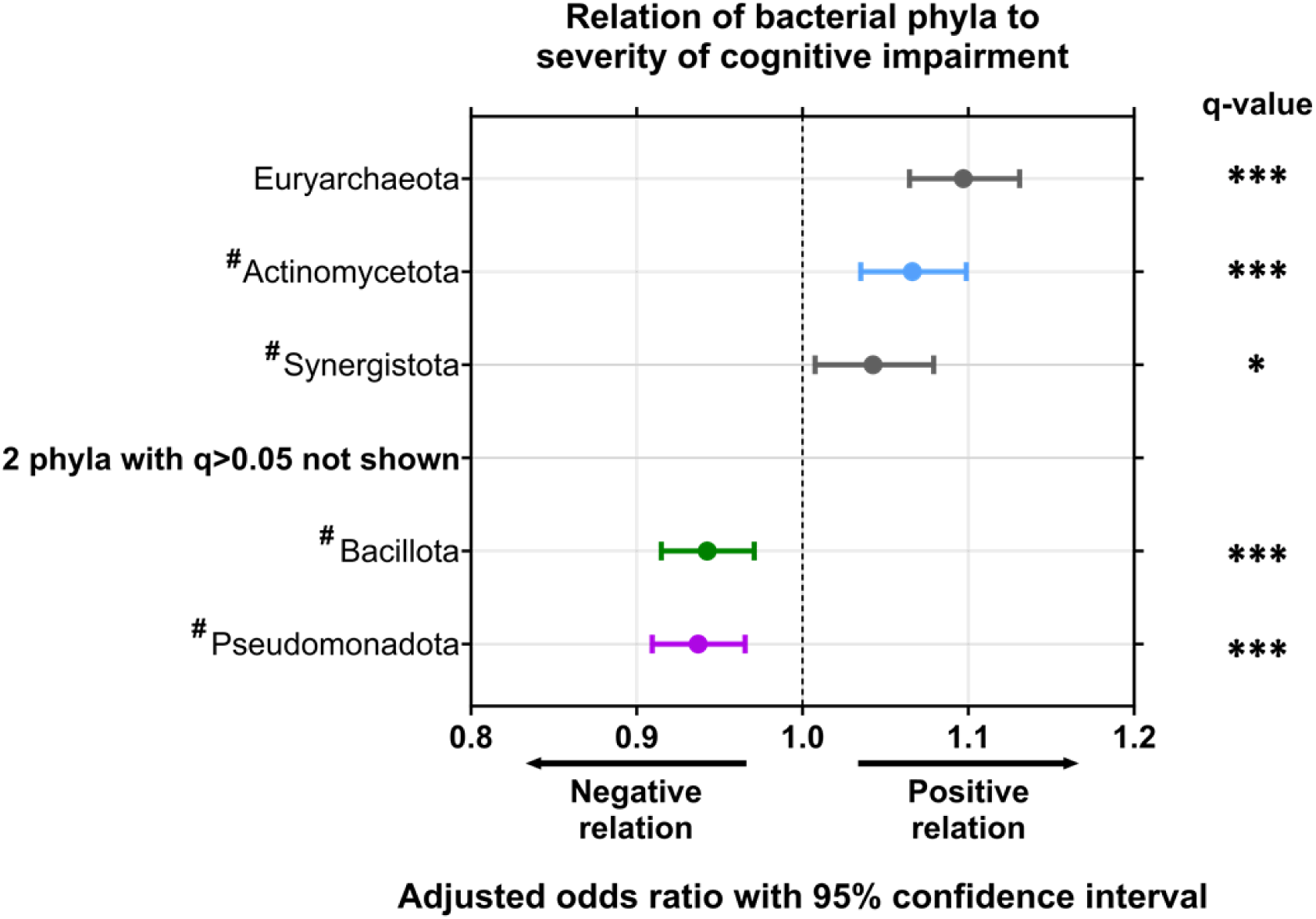
Phyla-level differences in the gut microbiome of residents of long-term aged care by cognitive impairment. Odds ratio and 95% confidence interval of effect of cognitive impairment severity on phyla relative abundance. Colours indicate bacterial phyla: blue = Actinomycetota; green = Bacillota; purple = Pseudomonadota; grey = non-core species. Performed by multivariate analysis, adjusting for time since cognitive impairment assessment, age, sex, antibiotic use, proton pump inhibitor use, opioid use, laxative use, recorded medical history, meal texture, and liquid texture. ^#^Denotes phyla with recently amended names: Actinomycetota (Actinobacteria), Synergistota (Synergistetes), Bacillota (Firmicutes), and Pseudomonadota (Proteobacteria). *q<0.05; ***q<0.001 for adjusted p-values following FDR correction.

Bacterial species that were detected in at least 60% of participants, and with a relative abundance of at least 0.1%, were denoted as “core” taxa (**Supplementary Table 2**). Of the 586 microbial species identified across the entire cohort, 30 were identified as core in mild CI (**Supplementary** Fig. 5A), 31 in moderate CI (**Supplementary** Fig. 5B), and 29 in severe CI (**Supplementary** Fig. 5C).

Comparison of species relative abundances identified 50 species that differed significantly with CI severity (**Fig. 3A**). Notably, *Blautia hydrogentrophica* (OR: 1.135 [1.099, 1.173]), *Catabacter hongkongensis* (OR: 1.131 [1.096, 1.168]), and *Alistipes finegoldii* (OR: 1.089 [1.058, 1.121]), had the strongest positive association with CI severity (all q<0.001, **Fig. 3A**). Further, *Collinsella aerofaciens* and *Methanobrevibacter smithii* were not only positively associated with CI severity (q<0.001, **Fig. 3A**), they were also core species in severe CI, but not mild or moderate (**Fig. 3B**, **Supplementary Table 2, Supplementary** Fig. 5). In contrast, *Bacteroides uniformis* (OR: 0.935 [0.908, 0.962]), *Blautia producta* (OR: 0.916 [0.888, 0.945]) and *Blautia wexlerae* (OR: 0.940 [0.913, 0.968]) were among those with the strongest inverse association with CI severity (all q<0.001, **Fig. 3A**). *Faecalibacterium prausnitzii*, a species previously associated with health outcomes in ageing (Jackson et al., 2016), was also found to trend lower in this cohort with increasing CI severity, but this did not achieve statistical significance (OR: 0.986 [0.958, 1.014], q=0.421, **Fig. 3A**; p>0.05, **Fig. 3B**).

**Figure 3.**
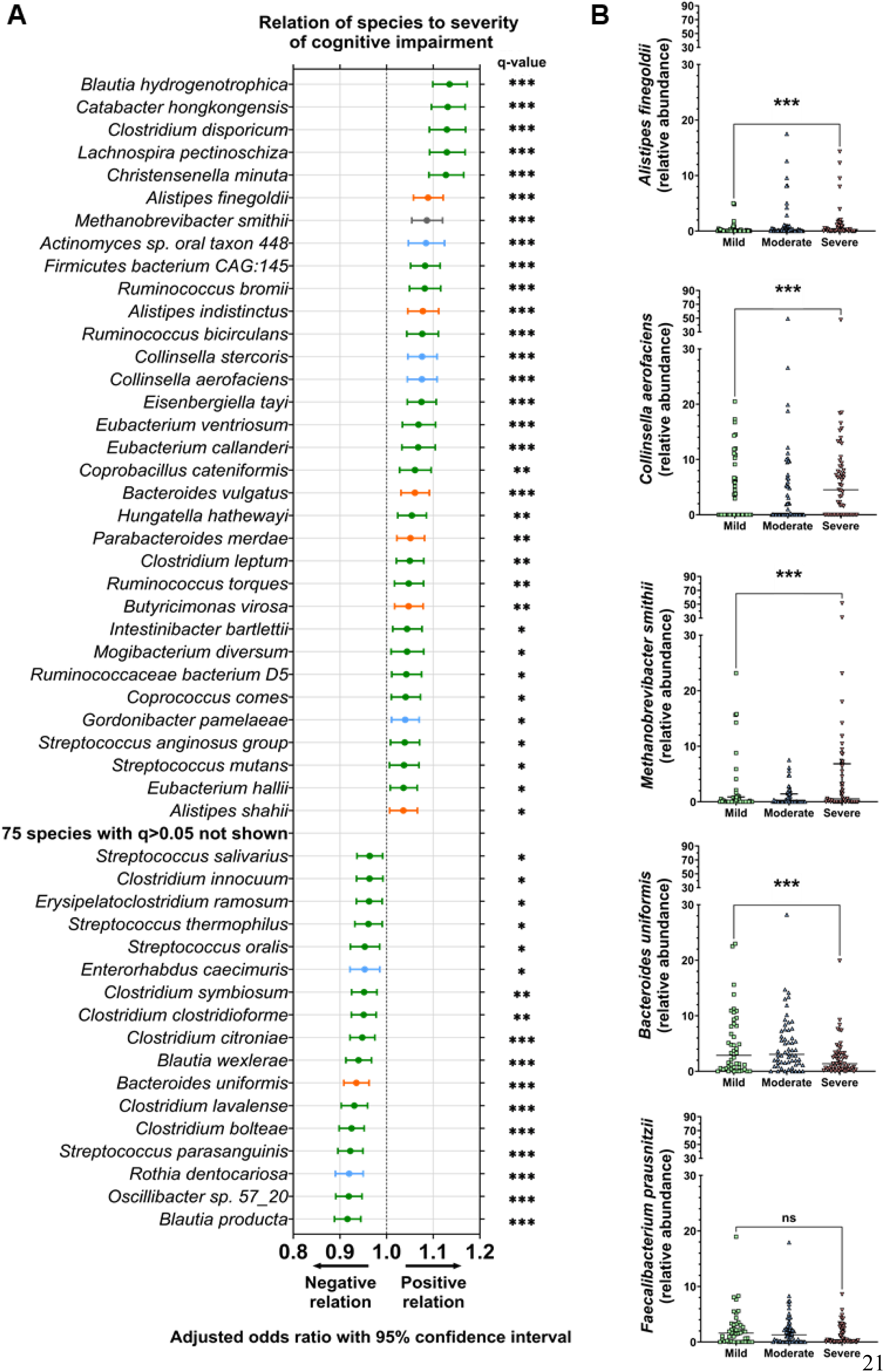
Species-level differences in the gut microbiome of residents of long-term aged care by cognitive impairment. **A**) Odds ratio and 95% confidence interval of effect of cognitive impairment severity on species relative abundance. Colours indicate bacterial phyla: blue = Actinomycetota; orange = Bacteroidota; green = Bacillota = green; grey = other. Performed by multivariate analysis, adjusting for time since cognitive impairment assessment, age, sex, antibiotic use, proton pump inhibitor use, opioid use, laxative use, recorded medical history, meal texture, and liquid texture. **B**) The relative abundance of bacterial species associated with cognitive impairment severity and aging: *Alistipes finegoldii*, *Collinsella aerofaciens*, *Methanobrevibacter smithii*, *Bacteroides uniformis*, and *Faecalibacterium prausnitzii*. ns=not significant; *q<0.05; **q<0.01; ***q<0.001 for adjusted p-values following FDR correction. Mild, n=46; moderate, n=58; severe, n=55.

### The functional capacity and output of the gut microbiota differs with CI severity

Differences in the functional capacity of the gut microbiota were identified with increasing CI severity. Four hundred MetaCyc pathways were detected in >30% of participants, of which, 70 were selected based on their potential influence on CI, including via mechanisms relating to neurotransmission, immunity, and metabolism. Metabolomic analysis of a subgroup of individuals (n=35; n=11-12/group) confirmed these findings (**Fig. 4A**). A total of 165 polar metabolites were detected in stool samples from these participants, including 33 metabolites classed as amino acids, peptides, and analogues, 50 classed as lipids and lipid-like molecules, 18 classed as carbohydrates, and 64 classed within other categories (**Supplementary** Fig. 6A), in addition to nine SCFAs (**Supplementary** Fig. 6B). Pathways inversely associated with CI severity included PWY-5505, a pathway essential to the production of the primary excitatory neurotransmitter glutamate (OR: 0.922 [0.895, 0.949], q<0.001), and GLUDEG-I-PWY, a pathway essential to the production of the primary inhibitory neurotransmitter GABA (OR: 0.962 [0.934, 0.990], q<0.05, **Fig. 4B**). The metabolite and excitatory neurotransmitter glutamate was present at lower levels in individuals with severe CI (**Fig. 4C**). Similarly, pathways related to the production of immunomodulatory SCFAs, including acetate (P461-PWY; OR: 0.877 [0.850, 0.904], q<0.001), propionate (P108-PWY; OR: 0.939 [0.913, 0.966], q<0.001), and butyrate (PWY-5022; OR: 0.935 [0.908, 0.962], q<0.001, **Fig. 4D**) were also lower in relative abundance as CI severity increased. The decrease in immune functional capacity corresponded with depleted levels of immunomodulating SCFA metabolites in individuals with severe CI, including for butyrate (q<0.01), propionate (q<0.01), and acetate (q<0.05, **Fig. 4E**; **Supplementary** Fig. 7). Functional pathways related to the biosynthesis of amino acids that regulate key metabolic processes, such as autophagy, included L-arginine (PWY-5154; OR: 0.911 [0.884, 0.938], q<0.001), L-lysine, L-threonine, and L-methionine (P4-PWY; OR: 0.905 [0.878, 0.932], q<0.001), and were among the most depleted at higher CI severity (**Fig. 4F**). Production of amino acid polar metabolites also decreased with increasing CI severity (**Fig. 4G**; **Supplementary** Fig. 7).

**Figure 4.**
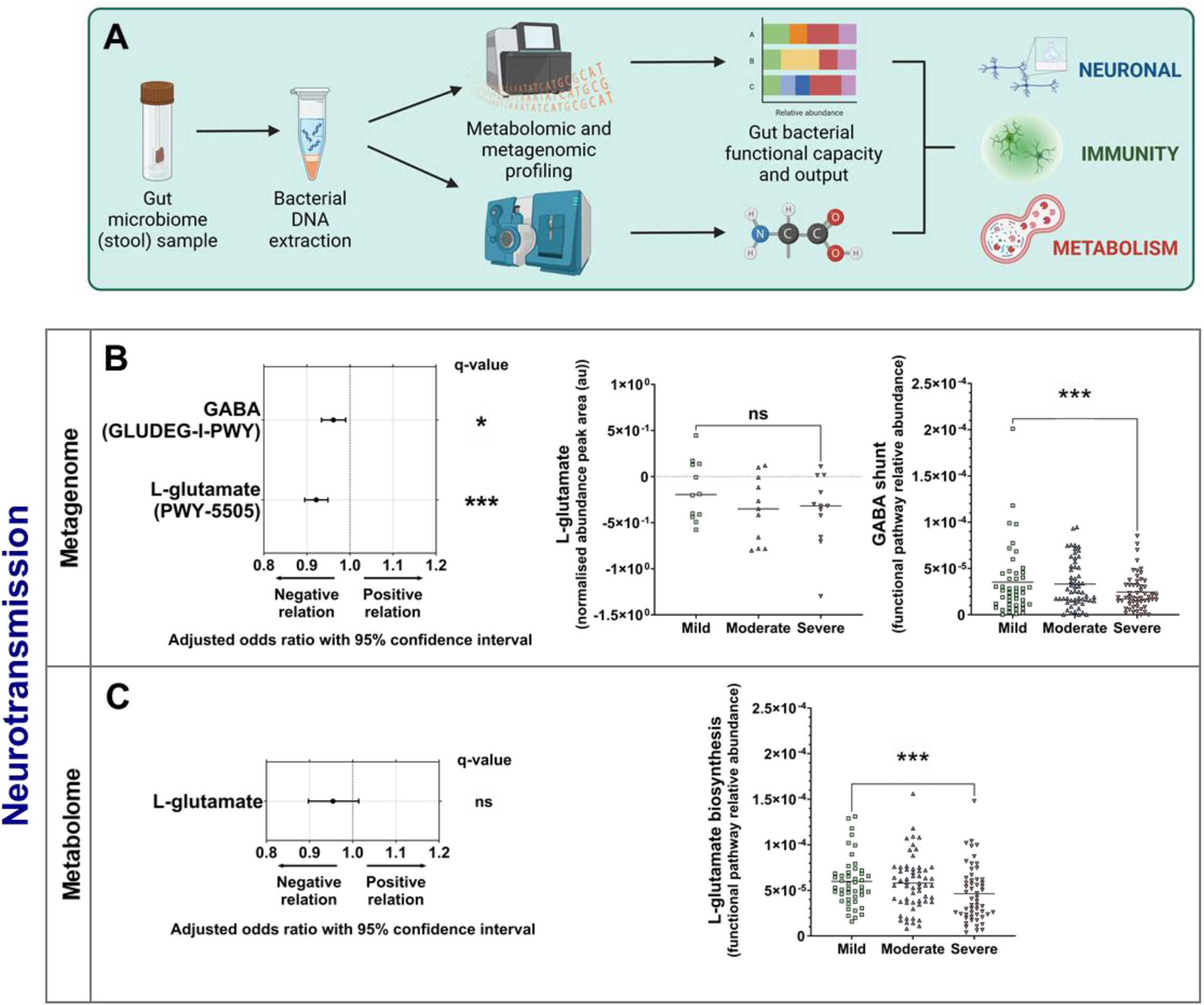
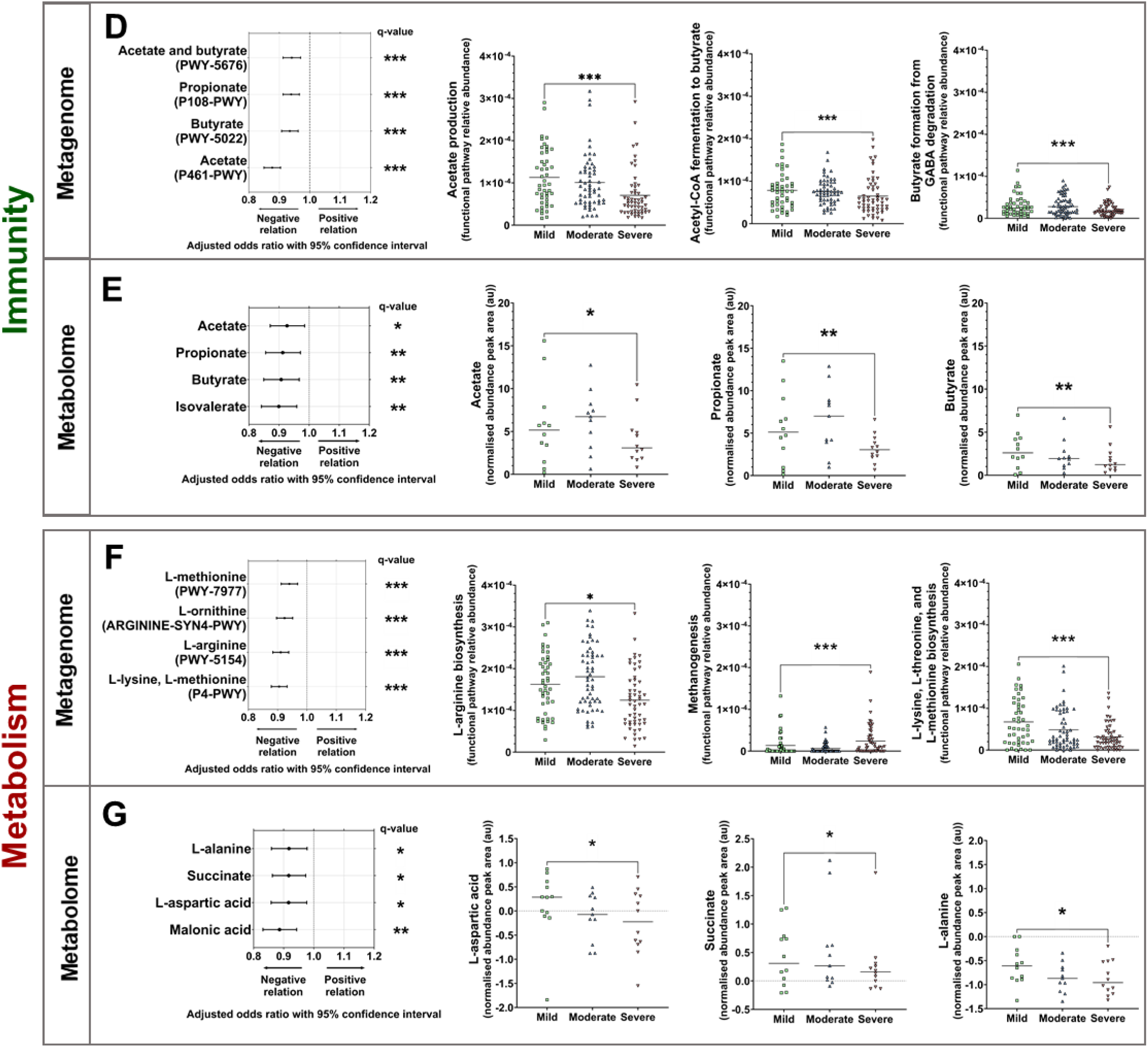
Specific functional differences relating to neurotransmission, immunity, and metabolism in the gut microbiome of residents of long-term aged care by cognitive impairment. **A**) Metagenomic and metabolomic profiling of microbiome functional capacity and output for long-term aged care residents with cognitive impairment in relation to neuronal communication (**B-C**), immunity (**D-E**), and metabolism (**F-G**). Odds ratio and 95% confidence interval of effect of cognitive impairment severity on functional pathway relative abundance and metabolite normalised abundance. Performed by multivariate analysis, adjusting for time since cognitive impairment assessment, age, sex, antibiotic use, proton pump inhibitor use, opioid use, laxative use, recorded medical history, meal texture, and liquid texture. The abundance of key pathways and metabolites grouped by cognitive impairment severity involved in neurotransmission, immunomodulation, and metabolism are shown. ns=not significant; *q<0.05; **q<0.01; ***q<0.001 for adjusted p-values following FDR correction. Pathways: mild, n=46; moderate, n=58; severe, n=55. Metabolites: mild, n=12; moderate, n=11; severe, n=12.

Further exploratory analysis across all functional pathways (n=400) identified 271 statistically significantly altered pathways, with multiple pathways related to methanogenesis among those of greatest significance and higher relative abundance (p<0.01, **Fig. 4F**; q<0.001, **Supplementary Table 3**). The relative abundances of metagenomic pathways and the levels of associated metabolites were also positively correlated (**Supplementary** Fig. 8).

## Discussion

We report significant associations between characteristics of the faecal microbiome and the severity of CI in residents of long-term aged care facilities. Microbiome CI severity-associated traits were identified even after adjustment for age, sex, prior medication exposure, and diet. Individuals with more severe CI exhibited a greater representation of the Actinomycetota phylum and *Methanobrevibacter smithii*, and a lower prevalence of *Bacteroides uniformis*, a reduced capacity for synthesis of SCFAs, neurotransmitters (glutamate and GABA), and amino acids that are essential for autophagy, and an increased capacity for methanogenesis. These findings identify microbial factors potentially influencing ageing-associated cognitive decline, and present opportunities for prediction and treatment of CI.

Changes in intestinal microbiology can influence neuroplastic changes in the brain via a range of mechanisms (Cryan et al., 2019; Rogers et al., 2016; Sharon et al., 2016; Shoubridge et al., 2022). Many of these pathways relate to the production of specific factors by the gut microbiota, including the biosynthesis of immunomodulatory metabolites and neurotransmitters (Correa-Oliveira et al., 2016; Erny et al., 2015), amino acid metabolism (Bellono et al., 2017; Ye et al., 2021), and the release of pro-inflammatory cytokines (Arentsen et al., 2017; Erny et al., 2015; Kim et al., 2013).

We assessed the potential influence of the gut microbiome of participants to neurophysiology through two complementary strategies. The first was the analysis of the metagenome, representing the functional capacity of microbes within the gut to produce particular metabolites. The second was a confirmatory analysis of the faecal metabolome, representing the output of the combined metabolic activity of the gut microbiota. Each of these processes identified factors that were significantly associated with CI severity, and notably, positive correlations between metabolite levels and the prevalence of genes involved in their biosynthesis was widespread.

A lower capacity for microbial biosynthesis of the neurotransmitters, glutamate and GABA, was evident in those with more severe CI. Levels of both factors have been associated with CI previously (Gueli & Taibi, 2013; Jimenez-Jimenez et al., 1998; Lin et al., 2019; Lin et al., 2017; Murley et al., 2020). The gut microbiome mediates neurological homeostasis via multiple key pathways, including through metabolism and production of neurotransmitters, such as glutamate, GABA, dopamine, and serotonin. These neurotransmitters can then directly innervate intestinal neural pathways or circulate peripherally to the brain (Kaelberer et al., 2018; Shoubridge et al., 2022; Strandwitz, 2018).

Severe CI was also associated with reduced capacity for bacterial biosynthesis of the SCFAs butyrate, acetate, and propionate. SCFA production is known to be important for normal cognitive function and in preventing neuroinflammation (Arnoldussen et al., 2017; Byrne et al., 2016; Erny et al., 2015). Previous studies have identified an association between a reduced capacity for SCFA biosynthesis and the development of a chronic and systemic inflammatory state, commonly referred to as “inflammaging”, involving increased circulation of IL-6, TNF-α, and C-reactive protein (Franceschi & Campisi, 2014; Franceschi et al., 2018; Frasca & Blomberg, 2016). Inflammaging, particularly in the brain, is associated with decreased neuronal arborisation, numbers of neurons and synapses, and overall brain cortical volume (Stephenson et al., 2018) and has been implicated in the acceleration of dementia onset (Grande et al., 2021; Sankowski et al., 2015), and the rate of neurological deterioration (Eikelenboom et al., 2012; Giunta et al., 2008).

In contrast to a decreased capacity for SCFA synthesis, we observed a greater capacity for methanogenesis with increasing CI severity. This relationship was apparent from the representation of methanogenic pathways within the metagenome, and from the increased relative abundance of species, such as *Methanobrevibacter smithii*, in those with severe CI. Increasing capacity for methanogenesis within the gut microbiome has been reported previously in two cohorts of centenarians (Li et al., 2022; Wu et al., 2019), as well as in rodent models of ageing (Maczulak et al., 1989). While the clinical consequences of increased methane production in the gut are poorly understood, high levels are associated with functional constipation (Chatterjee et al., 2007), diverticulosis (Weaver et al., 1986), and colon cancer (Haines et al., 1977).

The gut microbiome in participants with more severe CI was found to be depleted in its capacity to synthesise amino acids, particularly L-arginine. The availability of arginine is critical to the regulation of autophagy (Poillet-Perez et al., 2018), the cellular process that involves the recycling of nutrients from macromolecules in response to nutrient deficiency (Mizushima, 2004) and the removal of damaged material from the cellular environment (Lazarou et al., 2015). Genetic polymorphisms in genes involved in the regulation of autophagy have been linked to a number of neurodegenerative diseases, including Alzheimer’s, Parkinson’s, Huntington’s, and Lewy body diseases, frontotemporal dementia, and amyotrophic lateral sclerosis (Fujikake et al., 2018; Gan et al., 2018; Gao et al., 2018; Nixon, 2013; Tsuang et al., 2012). The conversion of arginine to putrescine, spermidine, and spermine by intestinal microbes promotes autophagy (Eisenberg et al., 2009; Oliphant & Allen-Vercoe, 2019; Pugin et al., 2017) and the significant reduction in arginine biosynthesis capacity is consistent with the contribution of suppressed autophagy to the development and progression of age-related disease. Severe CI was also associated with a reduced capacity for microbial production of the essential amino acids, L-valine and L-lysine. Impaired L-valine production has been linked with declining neurological health previously (Baranyi et al., 2016), and diet supplementation with L-lysine and L-valine has been shown to improve cognitive and psychological function in older adults (Suzuki et al., 2020).

Microbial functions, such as those associated with CI severity, can often be performed by many different members of the gut microbiota. This phenomenon is referred to as functional redundancy and can result in relationships between individual microbial species and host measures of disease being less strong than those based on conserved microbial functional traits. Despite this, we observed a number of discrete bacterial taxa that were significantly associated with CI. In particular, *Methanobrevibacter smithii* and *Alistipes finegoldii* were more prevalent in those with severe CI, while *Bacteroides uniformis* was less highly represented. As above, *Methanobrevibacter smithii* is associated with higher methane production (Ghoshal et al., 2016) and has been identified as an inflammatory and cardiometabolic biomarker (Fu et al., 2020). Whilst the precise mechanisms of *Alistipes* species in health and disease are still unclear (Parker et al., 2020), clinical studies of inflammatory diseases have shown *Alistipes finegoldii* triggers intestinal inflammation and decreases SCFA-producing bacteria, potentially playing a pathogenic role in chronic diseases (Kim et al., 2018; Parker et al., 2020; Rodriguez-Palacios et al., 2019).

We also observed severe CI to be associated with a lower prevalence of bacterial taxa that are considered broadly beneficial. These included *Bacteroides uniformis*, which is associated with reduced risk of colorectal cancer (Wang et al., 2012) and inflammatory bowel disease (Takahashi et al., 2016), and *Blautia* species, which have the potential to inhibit the growth of pathogenic bacteria in the intestine and reduce inflammation (Hosomi et al., 2022; Liu et al., 2021). Taxa that have been previously associated with aspects of cognition, such as *Collinsella aerofaciens* (Ghosh et al., 2020), were more prevalent in those with severe CI. However, other bacterial taxa associated with aspects of ageing, frailty, and cognitive decline in previous studies, including *Faecalibacterium prausnitzii* (Jackson et al., 2016), *Eubacterium rectale* (Ghosh et al., 2020; van Soest et al., 2020), and *Escherichia coli* (Barrientos et al., 2009; d’Avila et al., 2018; Hoogland et al., 2018) were not associated with CI severity in our cohort.

In addition to the relative abundance of different bacterial species, analysis of the broad structure of the gut microbiota can also be informative. We assessed three different alpha diversity measures, Shannon-Wiener diversity, Pielou’s evenness, and species richness, that together provide an overview of microbiota structure. While evenness, which represents the relative differences in the abundance of various species in the community, was not associated significantly with CI severity, Shannon-Wiener diversity and species richness were both higher in severe CI. Studies of the gut microbiota in ageing have previously reported reduced diversity in older age. For example, Verdi and colleagues identified significantly lower faecal microbiota diversity to be associated with longer reaction times (in cognitive assessments) in an independently living aged cohort (Verdi et al., 2018). Similarly, Wasser and colleagues reported reduced alpha diversity in those with Huntington’s disease (Wasser et al., 2020), while two other studies have reported no significant relationship between CI severity and alpha diversity (Komanduri et al., 2021; Stadlbauer et al., 2020). In our analysis, where age was adjusted for, a contrary effect was observed, consistent with an association between CI and specific microbiome changes that is separate to wider microbial shifts that are typical in later life.

Our study strengths and limitations include the use of entry into long term care facility assessment tools (‘ACFI’) for CI ascertainment. While it has been employed by the Australian Commonwealth Government as a basis for care funding for all residents of long term care facilities across Australia since 2008, is completed by trained assessors, and includes the PAS-CIS (a validated and consistently applied tool of CI in aged care), there are limitations to this tool. For example, people may not be comprehensively assessed for CI if they have a sensory/speech impairment, are non-English speaking, or have severe CI beyond the scope of the instrument, which can include a concurrent diagnosis of dementia or mental disorder (AIHW, 2002; Department of Health and Ageing, 2016). The ACFI is also designed for funding purposes, not clinical care or epidemiological surveillance, which likely results in underreporting of these chronic health conditions (Lind et al., 2020).

CI was measured on average 16 months prior to stool collection. There was no clinically significant difference in mean time from assessment to stool collection between severity groups, and the period of time between assessment and sample collection could not be reduced further without substantial reduction in the cohort size. However, changes in cognition between assessment and stool sample collection could confound the relationships that we report. Future studies arising from our findings would benefit from a briefer and more consistent interval between CI and microbiome assessment, and/or longitudinal analysis of associations.

Our study had other limitations that should be considered. First, we were able to relate changes in intestinal microbiology to cognitive function, but not to specific aspects of host neurological pathophysiology. Second, the relationships identified between taxonomic and functional features of the intestinal microbiome and CI are associative and whether they contribute directly to the development and progression of CI remains to be established. Third, the possibility that CI severity drives alterations in microbiome composition, mediated by factors such as dementia medications, changes in food preparation for those with dysphagia, and isolation to locked wards for residents with severe behavioural care needs, cannot be excluded based on the current analysis. Indeed, changes in behaviour associated with psychiatric conditions in other contexts, particularly those relating to diet, have been shown to contribute to disease-specific gut microbiome markers (Larroya et al., 2021).

While our analysis involved participants from five facilities in metropolitan South Australia, the findings are likely to be representative of a wider phenomenon. Alignment of the GRACE cohort to the comprehensive Registry of Senior Australians (ROSA) (Carpenter et al., 2023), a cohort that includes over 2.8 million Australians (including those in long-term aged care) supported the representative nature of our study cohort (Carpenter et al., 2023).

## Conclusions

We report age-, sex-, antibiotic-, and diet-independent microbial markers of severe CI. Our analysis implicates multiple gut microbiome-brain pathways in ageing-associated cognitive decline, including those involved in inflammation, neurotransmission, and autophagy. These findings raise the possibility of identifying cognitive decline and slowing its rate of progression via microbiome-targeted therapeutic interventions.

## DECLARATIONS

### Ethics approval and consent to participate

Ethical approval for the study was obtained from the Southern Adelaide Clinical Human Research Ethics Committee (HREC/18/SAC/244). Participants provided written informed consent themselves or where third-party consent was required, a legal guardian or family member with power of attorney provided consent on their behalf.

### Consent for publication

Not applicable.

### Data availability statement

The GRACE study data are available upon reasonable request. GRACE study data described in this article are available to all interested researchers through collaboration. Please contact GBR. The metagenomic data for this study have been deposited in the European Nucleotide Archive (ENA) at EMBL-EBI under accession number PRJEB51408 (https://www.ebi.ac.uk/ena/browser/view/PRJEB51408). The metabolomic data for this study is available at the NIH Common Fund’s National Metabolomics Data Repository (NMDR) website, the Metabolomics Workbench, https://www.metabolomicsworkbench.org (Sud et al., 2016), where it has been assigned Project ID PR001631. The metabolomic data can also be accessed directly via its Project DOI: http://dx.doi.org/10.21228/M8W43G. Regarding the sharing of linked data from ROSA, due to data custodian restrictions, individual ROSA data cannot be made publicly available to other researchers.

### Authors’ contributions

APS, LC, EF, LEP, MaC, MCI, KI, SLT, and GBR conceived and designed the study. APS, LC, EF, and JC acquired the clinical samples and clinical data. APS and LC acquired the metagenomic data. APS, DPDS and VKN acquired the metabolomic data. APS, LC, EF, JMC, KI, SLT, GBR analysed and interpreted the data. APS, LC, KI, SLT, and GBR drafted the manuscript. All authors provided intellectual input to the manuscript and critically revised the manuscript. All authors have read and approved the final version of the manuscript.

## Acknowledgments

This project used NCRIS-enabled Metabolomics Australia infrastructure at the University of Melbourne funded through BioPlatforms Australia. This project used the Metabolomics Workbench/National Metabolomics Data Repository (NMDR) supported by the NIH (U2C-DK119886) and Common Fund Data Ecosystem (CFDE) (3OT2OD030544).

