## Supplementary File for "Severe cognitive decline in long-term care is related to gut microbiome production of metabolites involved in neurotransmission, immunomodulation, and autophagy"

**SUPPLEMENTARY MATERIAL**

**Supplementary Text**

**Supplementary Text 1**

**Metabolite profiling methodology**

SCFA analysis was performed using an Agilent 6490 series triple quadrupole mass spectrometer (Agilent Technologies) with chromatographic separation on an Agilent 1200 series high-performance liquid chromatography system (Agilent Technologies). SCFAs were extracted by adding 360 μL of 50% acetonitrile with 10μM 4-methylvaleric acid internal standard to 40 μL of biological sample supernatant. Samples were then vortexed for 30 seconds, incubated at 10^o^C for 30 minutes at 950 rpm, centrifuged at 14,000 rpm for five minutes at 4^o^C, followed by supernatant collection. Derivatisation for SCFA analysis was performed by first adding 20 μL of 20 μM ^13^C_6_-nitrophenylhydrazine as internal standard to 40 μL of the extracted supernatant, followed by 20 μL each of 200 mM nitrophenylhydrazine and 120 mM 1-ethyl-3-(3-dimethylaminopropyl)carbodiimide (EDC), incubated at 40^o^C for 30 minutes at 950 rpm, quenched with 20 μL of 200 mM quinic acid, and incubated at 40^o^C for a further 30 minutes at 950 rpm. Lastly, the samples were reconstituted with 1.9 mL of 15% acetonitrile and 1 μL was injected onto the column. Pooled biological quality controls (PBQCs) were created by pooling extracts (20 μL) from individual biological samples and injected onto the column in five sample intervals. A reagent and procedural blank of the original sample preservation buffer was included for analysis to perform background correction.

Polar metabolite analysis was performed using an Agilent 6545 series quadrupole time-of-flight mass spectrometer (Agilent Technologies) with chromatographic separation on an Agilent 1200 series HPLC system (Agilent Technologies). Metabolite extraction was performed by first adding a solvent mixture of acetonitrile, methanol and water to 20 μL of biological sample, followed by vortexing, sonication, and agitation. Samples were centrifuged and supernatant collected and mixed with an internal standard mixture containing ^13^C_5_, ^15^N-valine, ^13^C_6_-leucine, and ^13^C_6_-sorbitol, and 14 μL of sample was injected onto the column. Samples were injected in a randomised order and PBQCs were injected onto the column in five sample intervals.

**Supplementary Tables and Figures**

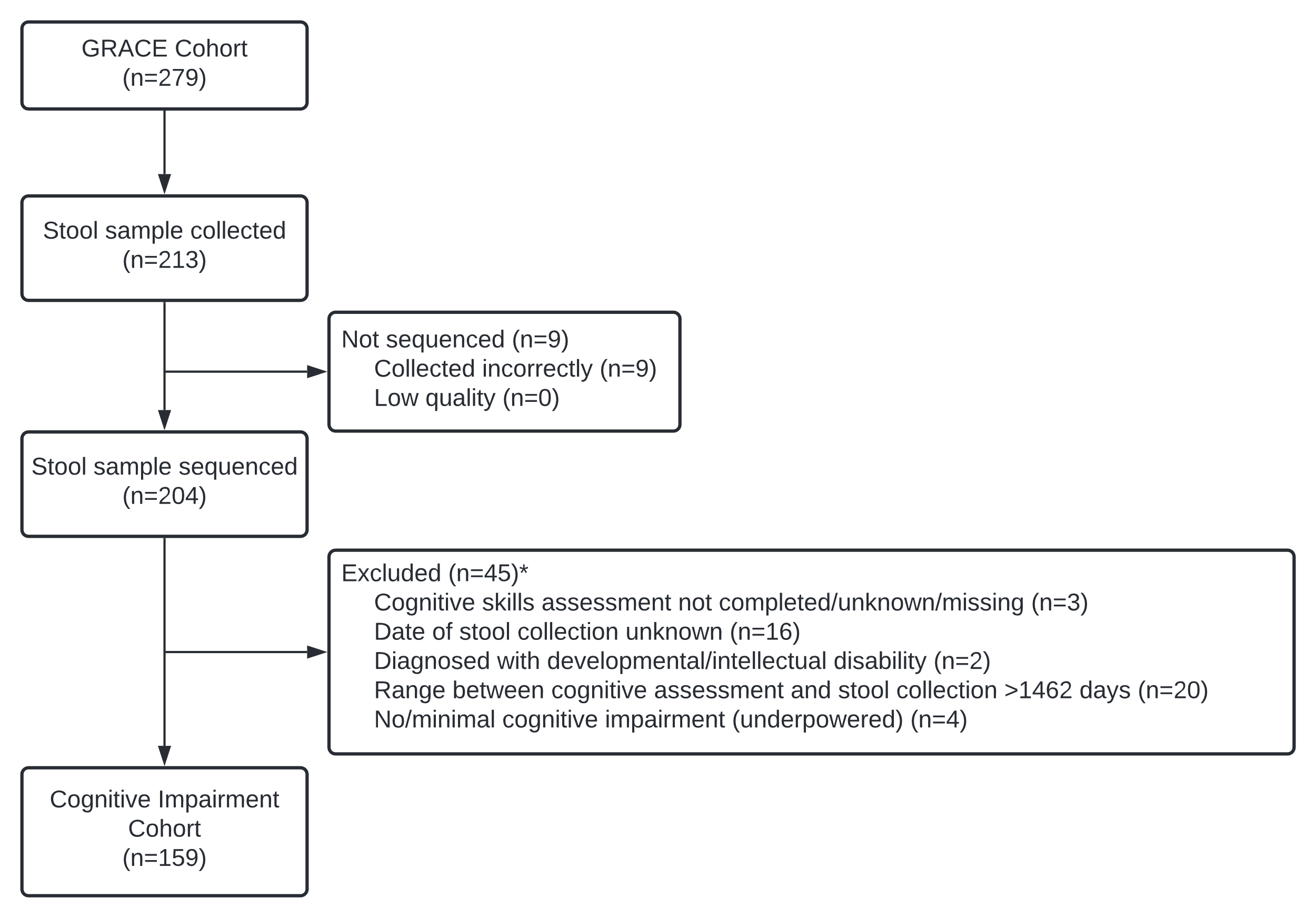

**Supplementary Figure 1. Selection of the Cognitive Impairment cohort.** Inclusion and exclusion numbers for the Cognitive Impairment cohort derived from the original GRACE cohort.

*Total number of participants excluded that met one or more of the exclusion criteria.

| Demographic | GRACE cohort  (n=279) | Cognitive Impairment cohort (n=159) |
| --- | --- | --- |
| Age (years):  Median (IQR) | 88.6 (81.8,93.2) | 88.6 (82.0,93.5) |
| Sex: % (n)  Female  Male | 71.7 (200)  28.3 (79) | 67.7 (109)  32.3 (52) |
| Time lived in facility (days):  Median (IQR) | 681 (252,1147) | 710 (361,1016) |
| Dementia diagnosis: % (n)* | 54.5 (152) | 56.6 (90) |
| Depression diagnosis: % (n)* | 56.6 (158) | 55.9 (90) |
| Delirium diagnosis: % (n)* | 5.7 (16) | 5.6 (9) |
| Cognitive Skills Rating: % (n)*^  Severe  Moderate  Mild  None or minimal | 28.0 (78)  39.8 (111)  27.6 (77)  2.9 (8) | 34.6 (55)  36.5 (58)  28.9 (46)  0.0 (0) |
| Activities of Daily Living care requirement: % (n)*^  High  Medium  Low | 65.9 (184)  26.5 (74)  6.5 (18) | 72.1 (116)  22.4 (36)  5.6 (9) |
| Cognition and Behaviour care requirement: % (n)*^  High  Medium  Low | 47.0 (131)  33.0 (92)  17.5 (49) | 46.0 (74)  38.5 (62)  15.5 (25) |
| Complex Healthcare care requirement: % (n)*^  High  Medium  Low | 64.5 (180)  28.3 (79)  6.1 (17) | 67.1 (108)  29.2 (47)  3.7 (6) |

Data are presented as median (IQR).

**Supplementary Table 1.** Cognitive Impairment cohort characteristics compared with the GRACE cohort.

*extracted from Aged Care Funding Instrument data.

^GRACE missing data: cognitive skills rating, 1.7% (n=5); activities of daily living care requirement, 1.1% (n=3); cognition and behaviour care requirement, 2.5% (n=7); complex healthcare care requirement, 1.1% (n=3); healthcare services, 12.9% (n=36); medications, 18.3% (n=51); health conditions, 18.3% (n=51).

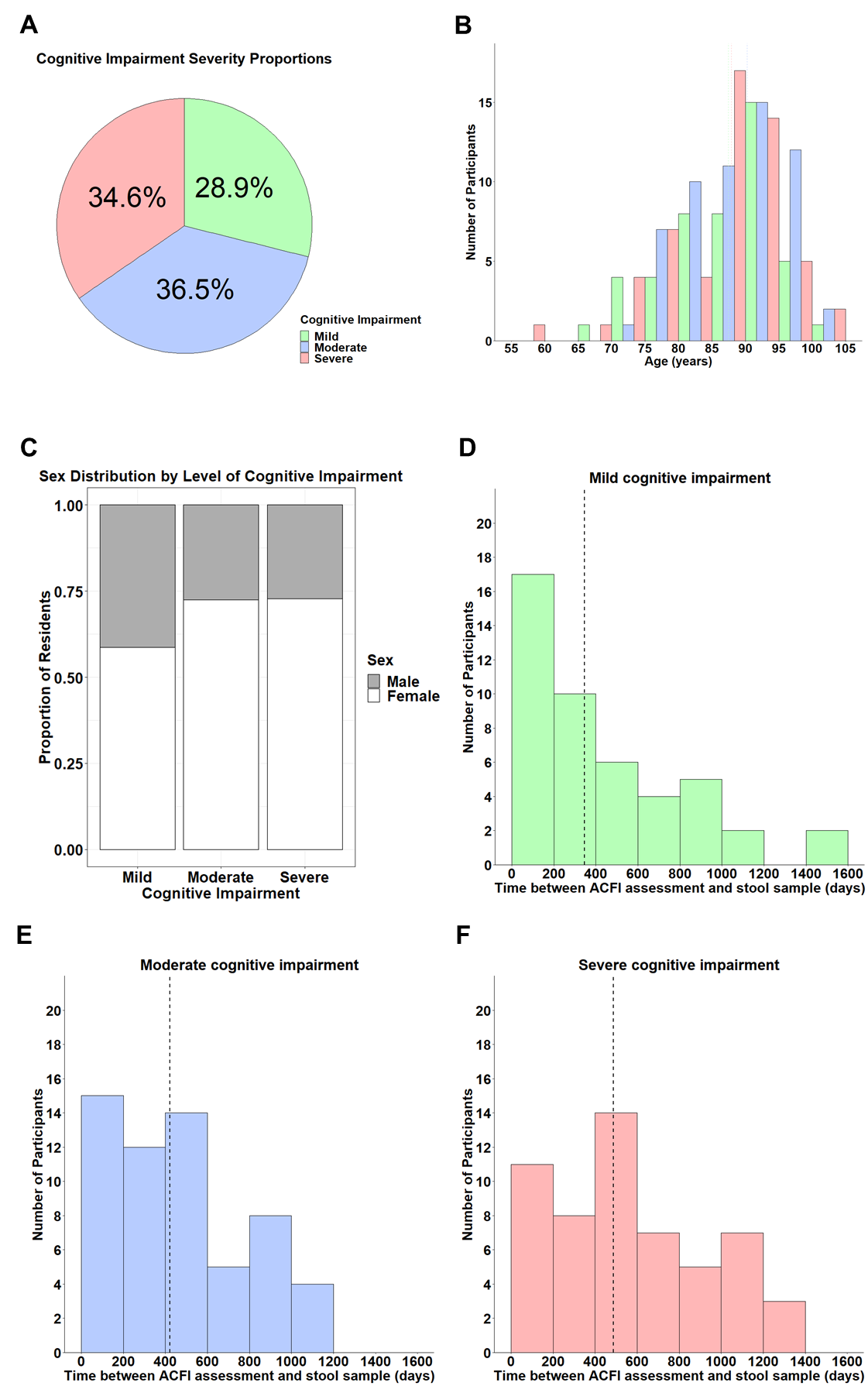

**Supplementary Figure 2. Characteristics of the cognitive impairment (CI) cohort.** **A**) The percentage of residents grouped by CI severity. The distribution of age (**B**) and sex (**C**) across CI groups within the CI cohort. The days between Cognitive Skills assessment and the collection of stool samples for mild (**D**), moderate (**E**), and severe (**F**) CI groups within the CI cohort. Mild, n=46; moderate, n=58; severe, n=55.

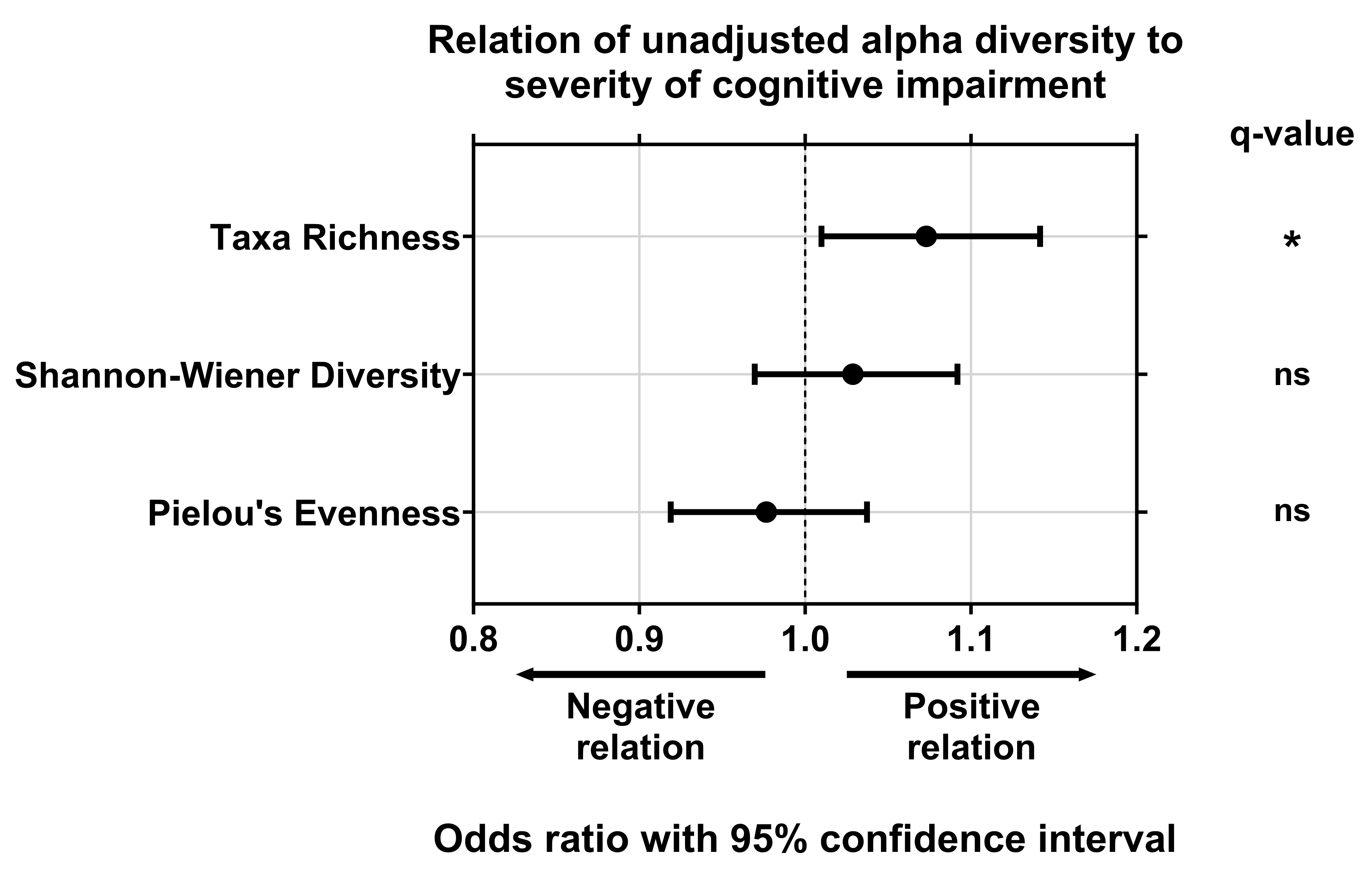

**Supplementary Figure 3. Alpha diversity of the gut microbiome of residents of long-term aged care by cognitive impairment.** Unadjusted odds ratio and 95% confidence interval of effect of cognitive impairment severity on microbiome diversity (taxa richness, Shannon-Wiener diversity, and Pielou’s evenness). ns=not significant; *q<0.05 for adjusted p-values following FDR correction.

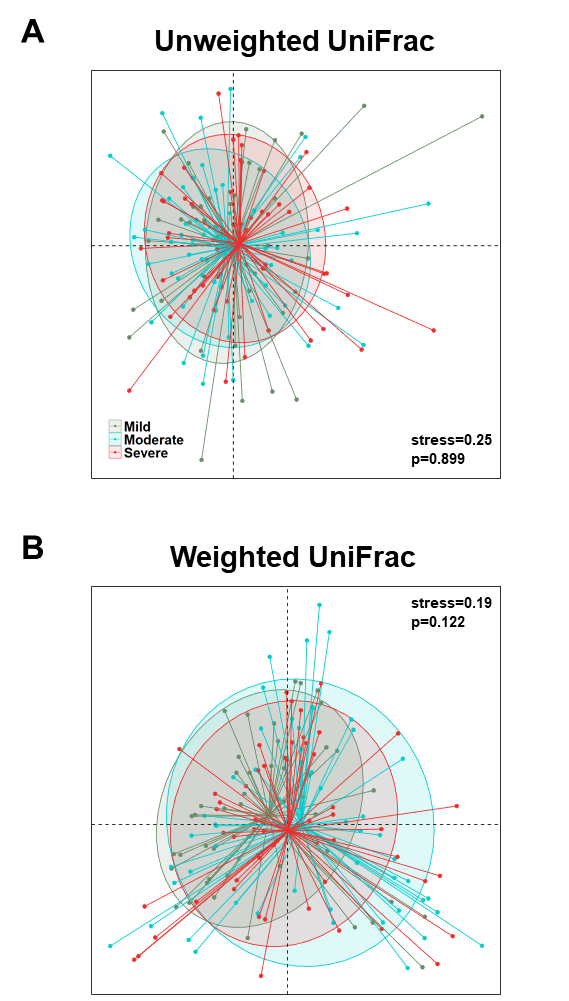

**Supplementary Figure 4. Non-metric multidimensional scaling plots of UniFrac dissimilarity of residents of long-term aged care facilities by cognitive impairment.** **A**) Unweighted UniFrac. **B**) Weighted UniFrac. P-values determined by unadjusted PERMANOVA. Cognitive impairment categorised as mild, n=46; moderate, n=58; severe, n=55.

| **Species name** | **Mild CI** | | **Moderate CI** | | **Severe CI** | |
| --- | --- | --- | --- | --- | --- | --- |
|  | **Prevalence (%)** | **Median abundance**  **% (range)** | **Prevalence (%)** | **Median abundance**  **% (range)** | **Prevalence (%)** | **Median abundance**  **% (range)** |
| *Eubacterium eligens* | 60.9 | 0.18 (0, 5.2) | - | - | 73.2 | 0.13 (0, 5.5) |
| *Eubacterium rectale* | 63.0 | 0.13 (0, 19.1) | - | - | 62.5 | 0.11 (0, 18.7) |
| *Bacteroides caccae* | - | - | 61.0 | 0.23 (0, 6.4) | - | - |
| *Akkermansia muciniphila* | - | - | 62.7 | 0.18 (0, 28.4) | - | - |
| *Blautia sp CAG 257* | - | - | 64.4 | 0.16 (0, 22.2) | - | - |
| *Firmicutes bacterium CAG 83* | - | - | 64.4 | 0.16 (0, 11.3) | 78.6 | 0.18 (0, 13.5) |
| *Dorea formicigenerans* | 63.0 | 0.24 (0, 4.7) | 66.1 | 0.20 (0, 6.3) | 83.9 | 0.28 (0, 3.9) |
| *Bacteroides vulgatus* | 63.0 | 0.29 (0, 13.4) | 69.5 | 1.05 (0, 31.4) | 83.9 | 1.40 (0, 10.3) |
| *Methanobrevibacter smithii* | - | - | - | - | 64.3 | 0.62 (0, 51.4) |
| *Roseburia faecis* | 65.2 | 0.11 (0, 8.7) | 64.4 | 0.19 (0, 30.9) | - | - |
| *Bacteroides thetaiotaomicron* | 67.4 | 0.10 (0, 8.7) | 84.7 | 0.43 (0, 12.2) | 85.7 | 0.11 (0, 5.3) |
| *Fusicatenibacter saccharivorans* | 67.4 | 0.61 (0, 12.3) | 76.3 | 0.58 (0, 8.4) | 67.9 | 0.25 (0, 11.2) |
| *Bacteroides dorei* | 67.4 | 0.69 (0, 22.6) | 62.7 | 0.40 (0, 13.2) | 76.8 | 0.29 (0, 5.2) |
| *Parabacteroides merdae* | - | - | 67.8 | 0.25 (0, 10.5) | 75.0 | 0.34 (0, 8.9) |
| *Eubacterium hallii* | 69.6 | 0.11 (0, 3.1) | 69.5 | 0.13 (0, 12.5) | 89.3 | 0.22 (0, 4.7) |
| *Odoribacter splanchnicus* | 69.6 | 0.13 (0, 1.3) | - | - | - | - |
| *Alistipes putredinis* | 69.6 | 1.03 (0, 7.4) | 74.6 | 0.76 (0, 4.8) | 78.6 | 0.61 (0, 6.1) |
| *Escherichia coli* | 71.7 | 0.20 (0, 16.5) | 79.7 | 0.39 (0, 43.6) | 83.9 | 0.23 (0, 19.6) |
| *Bifidobacterium longum* | 71.7 | 1.08 (0, 35.6) | 64.4 | 0.32 (0, 34.9) | 73.2 | 1.71 (0, 60.2) |
| *Collinsella aerofaciens* | - | - | - | - | 73.2 | 4.91 (0, 47.5) |
| *Firmicutes bacterium_CAG_83* | 73.9 | 0.35 (0, 9.6) | - | - | - | - |
| *Agathobaculum butyriciproducens* | - | - | 74.6 | 0.16 (0, 6.9) | - | - |
| *Anaerostipes hadrus* | 78.3 | 1.27 (0, 14.1) | 72.9 | 1.08 (0, 17.5) | 94.6 | 1.30 (0, 34.4) |
| *Alistipes finegoldii* | - | - | 76.3 | 0.21 (0, 17.5) | 82.1 | 0.26 (0, 14.3) |
| *Erysipelatoclostridium ramosum* | - | - | 79.7 | 0.10 (0, 9.1) | - | - |
| *Faecalibacterium prausnitzii* | 80.4 | 1.62 (0, 19.0) | 79.7 | 1.17 (0, 17.9) | 92.9 | 0.43 (0, 8.6) |
| *Blautia wexlerae* | 82.6 | 0.15 (0, 13.0) | 79.7 | 0.20 (0, 9.4) | - | - |
| *Streptococcus salivarius* | 82.6 | 0.33 (0, 7.3) | 81.4 | 0.18 (0, 12.7) | - | - |
| *Eubacterium siraeum* | 82.6 | 0.42 (0, 5.0) | 74.6 | 0.39 (0, 10.3) | 82.1 | 0.35 (0, 9.4) |
| *Blautia obeum* | 84.8 | 0.54 (0, 8.4) | 78.0 | 0.45 (0, 8.1) | 92.9 | 0.54 (0, 8.2) |
| *Parabacteroides distasonis* | 84.8 | 0.62 (0, 6.1) | 86.4 | 0.67 (0, 13.4) | 92.9 | 0.54 (0, 7.9) |
| *Ruminococcus gnavus* | 87.0 | 0.58 (0, 53.1) | 88.1 | 0.83 (0, 19.9) | 91.1 | 0.41 (0, 15.1) |
| *Streptococcus parasanguinis* | 89.1 | 0.11 (0, 4.6) | - | - | - | - |
| *Bacteroides uniformis* | 89.1 | 2.89 (0, 23.0) | 88.1 | 2.92 (0, 28.2) | 89.3 | 1.36 (0, 19.9) |
| *Clostridium leptum* | 91.3 | 0.11 (0, 1.4) | - | - | 96.4 | 0.17 (0, 1.9) |
| *Flavonifractor plautii* | 91.3 | 0.15 (0, 4.2) | 91.5 | 0.23 (0, 6.4) | 89.3 | 0.11 (0, 2.4) |
| *Clostridium innocuum* | 95.7 | 0.22 (0, 13.3) | - | - | 92.9 | 0.12 (0, 4.2) |
| *Eggerthella lenta* | 95.7 | 1.25 (0, 15.6) | 96.6 | 0.80 (0, 10.8) | 96.4 | 1.43 (0, 18.9) |
| *Gordonibacter pamelaeae* | 97.8 | 0.25 (0, 2.7) | 98.3 | 0.19 (0, 5.4) | 98.2 | 0.43 (0, 3.9) |
| *Ruthenibacterium lactatiformans* | 100.0 | 0.53 (0, 10.9) | 100.0 | 0.42 (0, 8.9) | 100.0 | 0.82 (0, 12.3) |

**Supplementary Table 2.** Prevalence and relative abundance of species identified as core (present in >60% of participants) in the gut microbiome of residents of long-term aged care facilities with mild, moderate, and severe cognitive impairment (CI). Taxa not identified in a CI group were below detected threshold. CI; cognitive impairment.

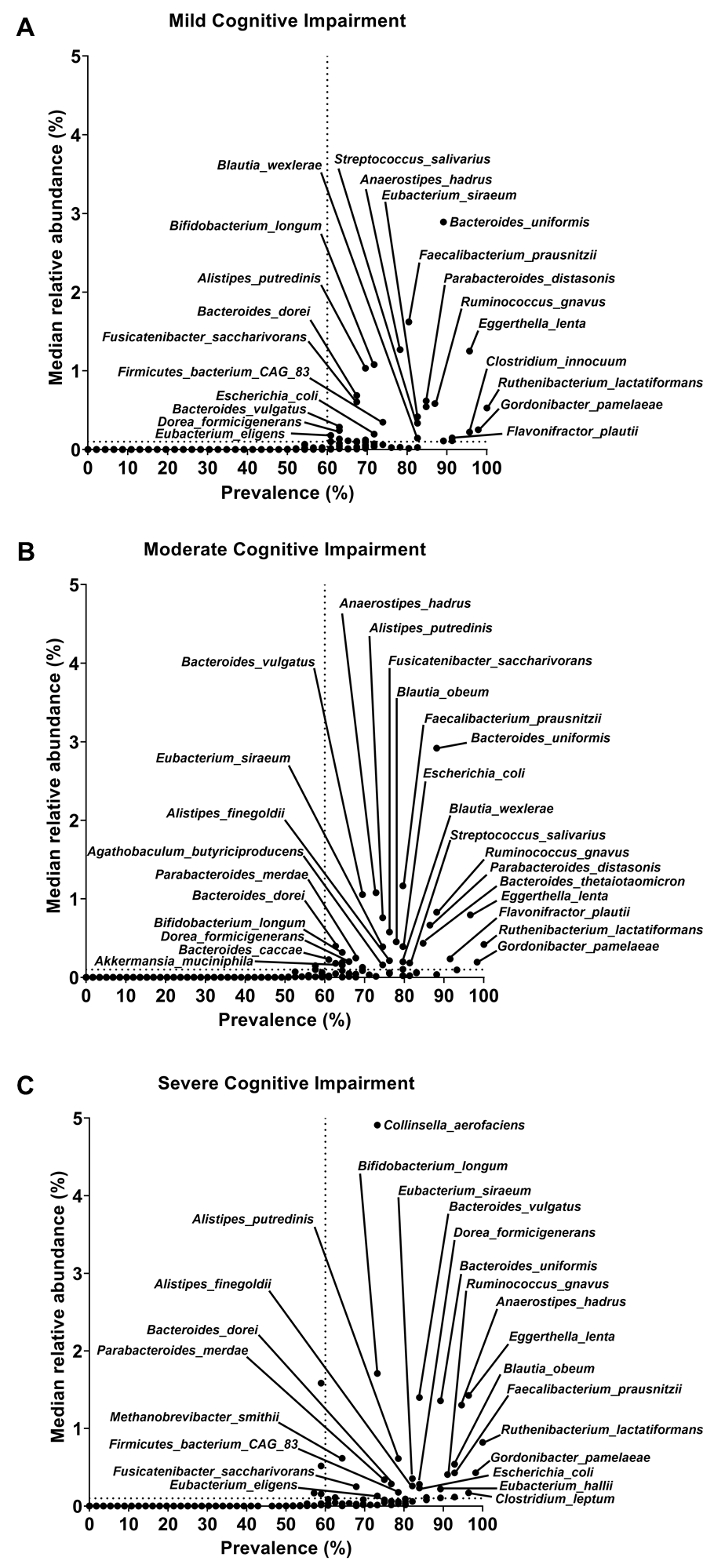

**Supplementary Figure 5. Core species profiles by cognitive impairment (CI) severity.** The frequency of species detected in the gut microbiome and their median relative abundances for: **A**) mild CI, **B**) moderate CI, and **C**) severe CI. Labelled species above core threshold.

| odds.ratio | c.int.low | c.int.high | pathway | p.value | fdr |
| --- | --- | --- | --- | --- | --- |
| 1.113094 | 1.078745838 | 1.149154726 | PWY.7286..7..3.amino.3.carboxypropyl..wyosine.biosynthesis | 2.99E-11 | 6.33E-10 |
| 1.107933 | 1.073984937 | 1.143529212 | PWY.8112..factor.420.biosynthesis.I..archaea. | 1.48E-10 | 2.20E-09 |
| 1.107342 | 1.073282852 | 1.143069752 | PWY.6350..archaetidylinositol.biosynthesis | 2.17E-10 | 3.11E-09 |
| 1.105943 | 1.071803128 | 1.141740534 | PWY.8113..3PG.factor.420.biosynthesis | 4.14E-10 | 5.04E-09 |
| 1.104314 | 1.070596543 | 1.139645541 | PWY.5198..factor.420.biosynthesis.II..mycobacteria. | 4.75E-10 | 5.62E-09 |
| 1.10393 | 1.068642914 | 1.141098788 | PWY.6167..flavin.biosynthesis.II..archaea. | 3.37E-09 | 2.60E-08 |
| 1.099599 | 1.06632844 | 1.134415866 | PWY.6349..CDP.archaeol.biosynthesis | 1.78E-09 | 1.60E-08 |
| 1.097014 | 1.064550378 | 1.130712389 | PWY.7198..pyrimidine.deoxyribonucleotides.de.novo.biosynthesis.IV | 1.61E-09 | 1.58E-08 |
| 1.096986 | 1.063753135 | 1.131746296 | PWY.6160..3.dehydroquinate.biosynthesis.II..archaea. | 4.62E-09 | 3.44E-08 |
| 1.096986 | 1.063753135 | 1.131746296 | PWY.6165..chorismate.biosynthesis.II..archaea. | 4.62E-09 | 3.44E-08 |
| 1.091733 | 1.058434844 | 1.126579998 | METHANOGENESIS.PWY..methanogenesis.from.H2.and.CO2 | 3.44E-08 | 2.13E-07 |
| 1.08915 | 1.055992566 | 1.123841604 | PWY.5209..methyl.coenzyme.M.oxidation.to.CO2 | 7.48E-08 | 3.80E-07 |
| 1.088497 | 1.055950561 | 1.122428967 | PWY.8187..L.arginine.degradation.XIII..reductive.Stickland.reaction. | 4.73E-08 | 2.72E-07 |
| 1.087369 | 1.051848025 | 1.124795631 | PWY.7159..3.8.divinyl.chlorophyllide.a.biosynthesis.III..aerobic..light.independent. | 9.56E-07 | 3.77E-06 |
| 1.084402 | 1.048690756 | 1.122071647 | PWY.7031..protein.N.glycosylation..bacterial. | 2.60E-06 | 9.24E-06 |
| 1.071246 | 1.037109738 | 1.107020183 | PWY.6518..bile.acids.epimerization | 3.49E-05 | 9.81E-05 |
| 1.063492 | 1.033363444 | 1.094611743 | PWY.7383..anaerobic.energy.metabolism..invertebrates..cytosol. | 2.80E-05 | 7.98E-05 |
| 1.059425 | 1.026396989 | 1.093948956 | PWY.5531..3.8.divinyl.chlorophyllide.a.biosynthesis.II..anaerobic. | 0.000381 | 0.000816 |
| 1.055535 | 1.024389794 | 1.087561131 | UDPNAGSYN.PWY..UDP.N.acetyl.D.glucosamine.biosynthesis.I | 0.000358 | 0.000778 |
| **1.048846** | **1.018965188** | **1.079828715** | **HISTSYN.PWY..L.histidine.biosynthesis** | **0.001238** | **0.002439** |
| 1.047155 | 1.017568965 | 1.077756687 | PWY.7209..superpathway.of.pyrimidine.ribonucleosides.degradation | 0.001662 | 0.003151 |
| **1.047085** | **1.016801803** | **1.078582718** | **PWY.2942..L.lysine.biosynthesis.III** | **0.002254** | **0.004118** |
| 1.046363 | 1.015822864 | 1.077824369 | PWY.1861..formaldehyde.assimilation.II..assimilatory.RuMP.Cycle. | 0.002573 | 0.004638 |
| 1.046028 | 1.016334804 | 1.07668889 | PWY66.399..gluconeogenesis.III | 0.002199 | 0.004037 |
| 1.043559 | 1.01408131 | 1.074014708 | RUMP.PWY..formaldehyde.oxidation.I | 0.003588 | 0.006298 |
| **1.043065** | **1.013395656** | **1.073679943** | **P124.PWY..Bifidobacterium.shunt** | **0.004213** | **0.007301** |
| 1.038445 | 1.008157506 | 1.069885067 | NONMEVIPP.PWY..methylerythritol.phosphate.pathway.I | 0.01286 | 0.020763 |
| 1.038307 | 1.008513155 | 1.069171527 | OANTIGEN.PWY..O.antigen.building.blocks.biosynthesis..E..coli. | 0.011886 | 0.019345 |
| 1.036371 | 1.007197838 | 1.066550144 | FASYN.INITIAL.PWY..superpathway.of.fatty.acid.biosynthesis.initiation..E..coli. | 0.014391 | 0.022866 |
| 1.033883 | 1.003744129 | 1.064470963 | PWY.5686..UMP.biosynthesis.I | 0.02524 | 0.038145 |
| 1.033883 | 1.003744129 | 1.064470963 | PWY.7790..UMP.biosynthesis.II | 0.02524 | 0.038145 |
| 1.033883 | 1.003744129 | 1.064470963 | PWY.7791..UMP.biosynthesis.III | 0.02524 | 0.038145 |
| 1.033566 | 1.004604078 | 1.063459744 | PWY.622..starch.biosynthesis | 0.022938 | 0.035466 |
| 1.032343 | 1.00311404 | 1.062512074 | PWY.6122..5.aminoimidazole.ribonucleotide.biosynthesis.II | 0.030636 | 0.045614 |
| 1.032343 | 1.00311404 | 1.062512074 | PWY.6277..superpathway.of.5.aminoimidazole.ribonucleotide.biosynthesis | 0.030636 | 0.045614 |
| 1.03142 | 1.001035129 | 1.062426454 | PWY0.162..superpathway.of.pyrimidine.ribonucleotides.de.novo.biosynthesis | 0.040879 | 0.059976 |
| 1.0289 | 1.000346374 | 1.05837268 | PWY.7371..1.4.dihydroxy.6.naphthoate.biosynthesis.II | 0.047511 | 0.069452 |
| 0.969588 | 0.941716645 | 0.998138128 | BIOTIN.BIOSYNTHESIS.PWY..biotin.biosynthesis.I | 0.03735 | 0.054999 |
| 0.969585 | 0.941921933 | 0.997876905 | PWY.7688..dTDP..alpha..D.ravidosamine.and.dTDP.4.acetyl..alpha..D.ravidosamine.biosynthesis | 0.035817 | 0.052935 |
| 0.969214 | 0.941398871 | 0.997459178 | ILEUSYN.PWY..L.isoleucine.biosynthesis.I..from.threonine. | 0.032565 | 0.048307 |
| 0.968041 | 0.940604831 | 0.996227949 | PWY0.42..2.methylcitrate.cycle.I | 0.026611 | 0.040067 |
| **0.967749** | **0.940363321** | **0.995628835** | **VALSYN.PWY..L.valine.biosynthesis** | **0.024757** | **0.037949** |
| 0.967703 | 0.939545278 | 0.996559304 | PWY.2941..L.lysine.biosynthesis.II | 0.028871 | 0.043306 |
| 0.967504 | 0.939900562 | 0.995767549 | PWY0.1479..tRNA.processing | 0.024827 | 0.037949 |
| 0.967242 | 0.939641574 | 0.995662934 | PWY.6124..inosine.5..phosphate.biosynthesis.II | 0.024252 | 0.037353 |
| 0.966689 | 0.940295831 | 0.993704318 | TCA..TCA.cycle.I..prokaryotic. | 0.016169 | 0.025391 |
| 0.966435 | 0.938374937 | 0.995156655 | PYRIDNUCSYN.PWY..NAD.de.novo.biosynthesis.I..from.aspartate. | 0.022329 | 0.034657 |
| 0.966329 | 0.938587333 | 0.994730975 | PWY.5913..partial.TCA.cycle..obligate.autotrophs. | 0.020915 | 0.032589 |
| 0.964887 | 0.937034347 | 0.993375047 | PWY.6317..D.galactose.degradation.I..Leloir.pathway. | 0.016353 | 0.02558 |
| 0.964486 | 0.937732766 | 0.991929347 | PWY.5464..superpathway.of.cytosolic.glycolysis..plants...pyruvate.dehydrogenase.and.TCA.cycle | 0.011598 | 0.018952 |
| 0.964435 | 0.936737231 | 0.992762791 | PWY.7560..methylerythritol.phosphate.pathway.II | 0.01433 | 0.02286 |
| 0.964054 | 0.935852852 | 0.99304867 | PWY0.461..L.lysine.degradation.I | 0.015492 | 0.024423 |
| 0.963519 | 0.935056575 | 0.992411331 | METH.ACETATE.PWY..methanogenesis.from.acetate | 0.014451 | 0.022871 |
| 0.963426 | 0.935512496 | 0.991780565 | PWY.5030..L.histidine.degradation.III | 0.011973 | 0.019408 |
| **0.962978** | **0.934421976** | **0.992233111** | **PWY.7400..L.arginine.biosynthesis.IV..archaebacteria.** | **0.013948** | **0.022339** |
| 0.962568 | 0.934827391 | 0.99087768 | GLYCOLYSIS.E.D..superpathway.of.glycolysis.and.the.Entner.Doudoroff.pathway | 0.010325 | 0.017011 |
| 0.962343 | 0.935705373 | 0.989597444 | PWY.5971..palmitate.biosynthesis..type.II.fatty.acid.synthase. | 0.007181 | 0.012128 |
| 0.962311 | 0.934723876 | 0.99044593 | PWY.8004..Entner.Doudoroff.pathway.I | 0.00936 | 0.015485 |
| 0.962182 | 0.933939058 | 0.991052114 | PWY.8131..5..deoxyadenosine.degradation.II | 0.010933 | 0.017938 |
| 0.962017 | 0.934376285 | 0.989983304 | PWY.5121..superpathway.of.geranylgeranyl.diphosphate.biosynthesis.II..via.MEP. | 0.009178 | 0.015246 |
| 0.961895 | 0.934741686 | 0.989673275 | METHGLYUT.PWY..superpathway.of.methylglyoxal.degradation | 0.007622 | 0.012768 |
| 0.961826 | 0.932533388 | 0.991952856 | PWY0.1277..3.phenylpropanoate.and.3..3.hydroxyphenyl.propanoate.degradation | 0.01344 | 0.021612 |
| **0.961622** | **0.933939652** | **0.990023986** | **GLUDEG.I.PWY..GABA.shunt** | **0.00849** | **0.014161** |
| 0.959655 | 0.932656756 | 0.987205741 | PWY.7210..pyrimidine.deoxyribonucleotides.biosynthesis.from.CTP | 0.00449 | 0.007649 |
| 0.958615 | 0.931140518 | 0.986706767 | GALACT.GLUCUROCAT.PWY..superpathway.of.hexuronide.and.hexuronate.degradation | 0.004236 | 0.007308 |
| **0.957995** | **0.930515044** | **0.985872943** | **HSERMETANA.PWY..L.methionine.biosynthesis.III** | **0.00375** | **0.006554** |
| 0.957931 | 0.930965176 | 0.985552988 | PWY.5705..allantoin.degradation.to.glyoxylate.III | 0.003091 | 0.005547 |
| 0.95781 | 0.930459441 | 0.985713768 | PWY.6588..pyruvate.fermentation.to.acetone | 0.00345 | 0.006136 |
| 0.957731 | 0.930293739 | 0.985933856 | PENTOSE.P.PWY..pentose.phosphate.pathway | 0.003515 | 0.006197 |
| 0.957721 | 0.927748097 | 0.988465074 | PWY0.41..allantoin.degradation.IV..anaerobic. | 0.007513 | 0.012638 |
| 0.957326 | 0.929264544 | 0.986125848 | PWY.5747..2.methylcitrate.cycle.II | 0.003976 | 0.00692 |
| 0.957121 | 0.928527455 | 0.985839714 | PWY.7456...beta...1.4..mannan.degradation | 0.003465 | 0.006136 |
| 0.957094 | 0.930216323 | 0.984632712 | PWY.5265..peptidoglycan.biosynthesis.II..staphylococci. | 0.002484 | 0.004499 |
| **0.956933** | **0.927582651** | **0.986969482** | **PWY.8190..L.glutamate.degradation.XI..reductive.Stickland.reaction.** | **0.005381** | **0.009127** |
| 0.956298 | 0.92959361 | 0.983608423 | PWY.6803..phosphatidylcholine.acyl.editing | 0.001916 | 0.003599 |
| 0.956264 | 0.927260814 | 0.986064367 | HCAMHPDEG.PWY..3.phenylpropanoate.and.3..3.hydroxyphenyl.propanoate.degradation.to.2.hydroxypentadienoate | 0.004326 | 0.007401 |
| 0.956264 | 0.927260814 | 0.986064367 | PWY.6690..cinnamate.and.3.hydroxycinnamate.degradation.to.2.hydroxypentadienoate | 0.004326 | 0.007401 |
| 0.956034 | 0.929808582 | 0.982871824 | TCA.GLYOX.BYPASS..superpathway.of.glyoxylate.bypass.and.TCA | 0.001487 | 0.002859 |
| 0.95588 | 0.928815746 | 0.983477673 | GALACTUROCAT.PWY..D.galacturonate.degradation.I | 0.002027 | 0.003773 |
| 0.95581 | 0.929463589 | 0.982754829 | GLYOXYLATE.BYPASS..glyoxylate.cycle | 0.001476 | 0.002853 |
| 0.955793 | 0.92819955 | 0.98387475 | PWY.6700..queuosine.biosynthesis.I..de.novo. | 0.002268 | 0.004125 |
| 0.955432 | 0.926733691 | 0.984739237 | PWY.7388..octanoyl..acyl.carrier.protein..biosynthesis..mitochondria..yeast. | 0.003219 | 0.005752 |
| 0.955409 | 0.928613424 | 0.982856921 | P221.PWY..octane.oxidation | 0.001626 | 0.003097 |
| 0.954984 | 0.927369188 | 0.98334059 | SALVADEHYPOX.PWY..adenosine.nucleotides.degradation.II | 0.002061 | 0.003818 |
| **0.954964** | **0.927383303** | **0.983157483** | **PWY.6292..superpathway.of.L.cysteine.biosynthesis..mammalian.** | **0.001969** | **0.003682** |
| 0.954719 | 0.927154018 | 0.982916107 | PWY1ZNC.1..assimilatory.sulfate.reduction.IV | 0.001858 | 0.003507 |
| **0.954621** | **0.927200818** | **0.9825786** | **CITRULBIO.PWY..L.citrulline.biosynthesis** | **0.001572** | **0.003009** |
| 0.954565 | 0.926570132 | 0.983092479 | PWY.7761..NAD.salvage.pathway.II..PNC.IV.cycle. | 0.002131 | 0.00393 |
| 0.953867 | 0.926536312 | 0.981798419 | PWY0.1241..ADP.L.glycero..beta..D.manno.heptose.biosynthesis | 0.001385 | 0.002704 |
| 0.953832 | 0.926361388 | 0.981961134 | PWY.7242..D.fructuronate.degradation | 0.001452 | 0.002819 |
| 0.953767 | 0.926520056 | 0.981634967 | REDCITCYC..TCA.cycle.VI..Helicobacter. | 0.001314 | 0.002577 |
| 0.952009 | 0.92466549 | 0.979954988 | PWY.6168..flavin.biosynthesis.III..fungi. | 0.000895 | 0.0018 |
| 0.951625 | 0.923655525 | 0.980163104 | PWY.6527..stachyose.degradation | 0.001084 | 0.002147 |
| 0.951438 | 0.923608105 | 0.980006655 | PWY.7237..myo...chiro..and.scyllo.inositol.degradation | 0.000988 | 0.001966 |
| 0.950764 | 0.923994321 | 0.978166781 | PWY.5675..nitrate.reduction.V..assimilatory. | 0.00051 | 0.001062 |
| 0.95047 | 0.92269037 | 0.978907233 | PWY4LZ.257..superpathway.of.fermentation..Chlamydomonas.reinhardtii. | 0.000755 | 0.001532 |
| **0.950333** | **0.922225424** | **0.97868403** | **PWY.702..L.methionine.biosynthesis.II** | **0.000712** | **0.001452** |
| 0.950286 | 0.923214861 | 0.9778794 | GLUCUROCAT.PWY..superpathway.of..beta..D.glucuronosides.degradation | 0.000516 | 0.00107 |
| 0.950253 | 0.924041188 | 0.977054175 | PWY.561..superpathway.of.glyoxylate.cycle.and.fatty.acid.degradation | 0.000334 | 0.000734 |
| 0.950074 | 0.922039912 | 0.978769243 | SULFATE.CYS.PWY..superpathway.of.sulfate.assimilation.and.cysteine.biosynthesis | 0.000767 | 0.00155 |
| 0.950041 | 0.922673568 | 0.978056242 | P125.PWY..superpathway.of..R.R..butanediol.biosynthesis | 0.000564 | 0.001163 |
| 0.94947 | 0.922343531 | 0.977169419 | PWY.241..C4.photosynthetic.carbon.assimilation.cycle..NADP.ME.type | 0.000449 | 0.000939 |
| 0.949432 | 0.922516972 | 0.976948167 | PWY.5918..superpathway.of.heme.b.biosynthesis.from.glutamate | 0.000385 | 0.000816 |
| 0.949173 | 0.922190939 | 0.976786006 | P23.PWY..reductive.TCA.cycle.I | 0.000375 | 0.000811 |
| 0.948887 | 0.922108087 | 0.976240472 | PWY.7184..pyrimidine.deoxyribonucleotides.de.novo.biosynthesis.I | 0.00031 | 0.000688 |
| 0.948884 | 0.92181819 | 0.976562705 | PWY.4041...gamma..glutamyl.cycle | 0.000357 | 0.000778 |
| 0.948686 | 0.921374227 | 0.976596912 | PWY.7315..dTDP.N.acetylthomosamine.biosynthesis | 0.000386 | 0.000816 |
| **0.948678** | **0.922207516** | **0.97573389** | **PWY.6922..L.N.delta..acetylornithine.biosynthesis** | **0.00025** | **0.000564** |
| **0.947645** | **0.919701081** | **0.976237995** | **PWY.5345..superpathway.of.L.methionine.biosynthesis..by.sulfhydrylation.** | **0.000407** | **0.000857** |
| 0.94753 | 0.919640745 | 0.976064915 | SO4ASSIM.PWY..assimilatory.sulfate.reduction.I | 0.000385 | 0.000816 |
| 0.947489 | 0.919784469 | 0.975335829 | CALVIN.PWY..Calvin.Benson.Bassham.cycle | 0.000271 | 0.000608 |
| 0.947377 | 0.920347257 | 0.974971456 | PWY.5189..tetrapyrrole.biosynthesis.II..from.glycine. | 0.000236 | 0.000535 |
| 0.947289 | 0.921077445 | 0.974093064 | AST.PWY..L.arginine.degradation.II..AST.pathway. | 0.000148 | 0.000353 |
| 0.946934 | 0.918685014 | 0.975342264 | PWY.6606..guanosine.nucleotides.degradation.II | 0.000316 | 0.000699 |
| 0.946646 | 0.91700856 | 0.976876176 | PWY66.430..myristate.biosynthesis..mitochondria. | 0.00067 | 0.001374 |
| 0.94662 | 0.919974632 | 0.973875998 | PWY.5723..Rubisco.shunt | 0.000158 | 0.000369 |
| 0.946275 | 0.918640919 | 0.974078803 | THISYNARA.PWY..superpathway.of.thiamine.diphosphate.biosynthesis.III..eukaryotes. | 0.000202 | 0.000463 |
| 0.946088 | 0.915348033 | 0.97761391 | PWY.7434..terminal.O.glycans.residues.modification..via.type.2.precursor.disaccharide. | 0.000953 | 0.001905 |
| 0.946055 | 0.919390128 | 0.973319831 | PWY.6284..superpathway.of.unsaturated.fatty.acids.biosynthesis..E..coli. | 0.000136 | 0.000333 |
| 0.945802 | 0.919391981 | 0.972759819 | PWY.5850..superpathway.of.menaquinol.6.biosynthesis | 0.000107 | 0.00027 |
| 0.945802 | 0.919391981 | 0.972759819 | PWY.5896..superpathway.of.menaquinol.10.biosynthesis | 0.000107 | 0.00027 |
| 0.945729 | 0.91803284 | 0.974058179 | GLYCOL.GLYOXDEG.PWY..superpathway.of.glycol.metabolism.and.degradation | 0.00022 | 0.000503 |
| 0.945726 | 0.918825408 | 0.973211502 | PWY.6906..chitin.derivatives.degradation | 0.000141 | 0.000344 |
| 0.945655 | 0.918565167 | 0.973346404 | PWY.6902..chitin.degradation.II..Vibrio. | 0.000161 | 0.000375 |
| 0.945328 | 0.91848974 | 0.972732472 | PWY.6545..pyrimidine.deoxyribonucleotides.de.novo.biosynthesis.III | 0.000122 | 0.000303 |
| 0.945248 | 0.916787691 | 0.97442452 | THREOCAT.PWY..superpathway.of.L.threonine.metabolism | 0.000292 | 0.000652 |
| 0.945215 | 0.917860168 | 0.97314409 | PWY.5497..purine.nucleobases.degradation.II..anaerobic. | 0.000158 | 0.000369 |
| 0.94517 | 0.917962312 | 0.97285609 | PWY.1042..glycolysis.IV | 0.000144 | 0.000346 |
| **0.94516** | **0.917569927** | **0.973116246** | **PWY.I9..L.cysteine.biosynthesis.VI..from.L.methionine.** | **0.000173** | **0.0004** |
| 0.944253 | 0.91663812 | 0.972427842 | PWY.6353..purine.nucleotides.degradation.II..aerobic. | 0.00015 | 0.000357 |
| **0.944025** | **0.916441237** | **0.972228963** | **P161.PWY..acetylene.degradation..anaerobic.** | **0.000132** | **0.000325** |
| 0.943997 | 0.915832558 | 0.972469539 | P185.PWY..formaldehyde.assimilation.III..dihydroxyacetone.cycle. | 0.000152 | 0.00036 |
| 0.943912 | 0.916328386 | 0.972110332 | PWY.7392..taxadiene.biosynthesis..engineered. | 0.000128 | 0.000316 |
| **0.943251** | **0.916620812** | **0.970478283** | **PWY.821..superpathway.of.sulfur.amino.acid.biosynthesis..Saccharomyces.cerevisiae.** | **5.99E-05** | **0.000156** |
| 0.94321 | 0.915971377 | 0.97097145 | PWY.5973..cis.vaccenate.biosynthesis | 8.73E-05 | 0.000224 |
| 0.942931 | 0.91655869 | 0.969842849 | PWY.5860..superpathway.of.demethylmenaquinol.6.biosynthesis.I | 4.53E-05 | 0.000123 |
| 0.942711 | 0.916361056 | 0.969630914 | PWY.6285..superpathway.of.fatty.acids.biosynthesis..E..coli. | 4.23E-05 | 0.000116 |
| 0.942648 | 0.915941397 | 0.969966317 | PWY.5138..fatty.acid..beta..oxidation.IV..unsaturated..even.number. | 5.30E-05 | 0.000141 |
| 0.942453 | 0.913985891 | 0.971584148 | PWY.7616..methanol.oxidation.to.carbon.dioxide | 0.000142 | 0.000344 |
| 0.942079 | 0.915283109 | 0.969505821 | FAO.PWY..fatty.acid..beta..oxidation.I..generic. | 4.79E-05 | 0.000129 |
| 0.941775 | 0.914588551 | 0.969574739 | PWY.5392..reductive.TCA.cycle.II | 5.57E-05 | 0.000147 |
| 0.94175 | 0.914256648 | 0.969754004 | POLYAMSYN.PWY..superpathway.of.polyamine.biosynthesis.I | 6.70E-05 | 0.000174 |
| 0.941357 | 0.913595347 | 0.969663734 | PWY.5384..sucrose.degradation.IV..sucrose.phosphorylase. | 7.04E-05 | 0.000181 |
| **0.940805** | **0.912435385** | **0.96975918** | **PWY.5676..acetyl.CoA.fermentation.to.butanoate.II** | **8.79E-05** | **0.000224** |
| 0.940313 | 0.912331392 | 0.968877235 | GOLPDLCAT.PWY..superpathway.of.glycerol.degradation.to.1.3.propanediol | 5.73E-05 | 0.00015 |
| 0.940221 | 0.912729308 | 0.968332509 | PWY.7345..superpathway.of.anaerobic.sucrose.degradation | 4.37E-05 | 0.000119 |
| **0.939994** | **0.912178778** | **0.968197666** | **PWY.7977..L.methionine.biosynthesis.IV** | **4.81E-05** | **0.000129** |
| 0.93994 | 0.912910862 | 0.967561407 | GLUCONEO.PWY..gluconeogenesis.I | 3.12E-05 | 8.84E-05 |
| 0.939703 | 0.912090683 | 0.967993604 | PWY.5855..ubiquinol.7.biosynthesis..early.decarboxylation. | 4.12E-05 | 0.000114 |
| 0.939012 | 0.912599114 | 0.965949519 | ORNDEG.PWY..superpathway.of.ornithine.degradation | 1.40E-05 | 4.17E-05 |
| **0.938887** | **0.912532986** | **0.965770297** | **P108.PWY..pyruvate.fermentation.to.propanoate.I** | **1.29E-05** | **3.90E-05** |
| 0.938824 | 0.912314936 | 0.965888267 | PWY0.1338..polymyxin.resistance | 1.43E-05 | 4.23E-05 |
| 0.938816 | 0.91071736 | 0.967171846 | PWY.6609..adenine.and.adenosine.salvage.III | 3.53E-05 | 9.85E-05 |
| 0.938433 | 0.911753531 | 0.965715722 | PWY.7118..chitin.deacetylation | 1.46E-05 | 4.29E-05 |
| 0.938359 | 0.91159511 | 0.965989399 | PWY.6895..superpathway.of.thiamine.diphosphate.biosynthesis.II | 1.67E-05 | 4.83E-05 |
| 0.93718 | 0.909735798 | 0.965138972 | PWY66.367..ketogenesis | 1.67E-05 | 4.83E-05 |
| 0.937168 | 0.910327546 | 0.964478632 | PWY.5695..inosine.5..phosphate.degradation | 1.04E-05 | 3.20E-05 |
| 0.936919 | 0.909371994 | 0.964962731 | PWY.7111..pyruvate.fermentation.to.isobutanol..engineered. | 1.75E-05 | 5.03E-05 |
| **0.936453** | **0.909204334** | **0.963825522** | **ASPASN.PWY..superpathway.of.L.aspartate.and.L.asparagine.biosynthesis** | **9.08E-06** | **2.85E-05** |
| 0.936426 | 0.909691637 | 0.963662948 | DENOVOPURINE2.PWY..superpathway.of.purine.nucleotides.de.novo.biosynthesis.II | 7.82E-06 | 2.56E-05 |
| 0.936039 | 0.908893847 | 0.963824358 | PWY.6897..thiamine.diphosphate.salvage.II | 9.97E-06 | 3.08E-05 |
| 0.93552 | 0.907935756 | 0.963610846 | PWY.6147..6.hydroxymethyl.dihydropterin.diphosphate.biosynthesis.I | 9.95E-06 | 3.08E-05 |
| 0.9355 | 0.90833836 | 0.963169708 | NAGLIPASYN.PWY..lipid.IVA.biosynthesis..E..coli. | 8.13E-06 | 2.61E-05 |
| 0.9355 | 0.90833836 | 0.963169708 | PWY.8073..lipid.IVA.biosynthesis..P..putida. | 8.13E-06 | 2.61E-05 |
| 0.935346 | 0.907465359 | 0.963765996 | PWY0.1296..purine.ribonucleosides.degradation | 1.37E-05 | 4.11E-05 |
| 0.935273 | 0.907654649 | 0.963370474 | PWY.6731..starch.degradation.III | 1.09E-05 | 3.31E-05 |
| 0.935033 | 0.908084077 | 0.962568914 | PWY.5367..petroselinate.biosynthesis | 6.15E-06 | 2.05E-05 |
| **0.934567** | **0.90767576** | **0.962031568** | **PWY.5022..4.aminobutanoate.degradation.V** | **5.03E-06** | **1.73E-05** |
| 0.934491 | 0.907999631 | 0.961561623 | PWY0.1337..oleate..beta..oxidation | 3.54E-06 | 1.24E-05 |
| 0.934257 | 0.906596754 | 0.962398837 | ARG.POLYAMINE.SYN..superpathway.of.arginine.and.polyamine.biosynthesis | 6.96E-06 | 2.29E-05 |
| 0.934144 | 0.906846541 | 0.96198656 | PWY.6703..preQ0.biosynthesis | 6.11E-06 | 2.05E-05 |
| 0.933788 | 0.907511929 | 0.96062417 | PWY.7942..5.oxo.L.proline.metabolism | 2.32E-06 | 8.31E-06 |
| 0.933711 | 0.90583812 | 0.962244701 | PWY0.1319..CDP.diacylglycerol.biosynthesis.II | 8.29E-06 | 2.62E-05 |
| 0.933711 | 0.90583812 | 0.962244701 | PWY.5667..CDP.diacylglycerol.biosynthesis.I | 8.29E-06 | 2.62E-05 |
| 0.933506 | 0.906855271 | 0.960652176 | PWY0.845..superpathway.of.pyridoxal.5..phosphate.biosynthesis.and.salvage | 2.83E-06 | 9.97E-06 |
| 0.933043 | 0.905873339 | 0.960638963 | PWY.6507..4.deoxy.L.threo.hex.4.enopyranuronate.degradation | 3.62E-06 | 1.26E-05 |
| 0.932728 | 0.906240366 | 0.959805671 | PWY.7094..fatty.acid.salvage | 1.97E-06 | 7.13E-06 |
| 0.932037 | 0.906202138 | 0.958385953 | PWY.7858...5Z..dodecenoate.biosynthesis.II | 8.21E-07 | 3.32E-06 |
| 0.931269 | 0.904485575 | 0.958583893 | NAD.BIOSYNTHESIS.II..NAD.salvage.pathway.III..to.nicotinamide.riboside. | 1.53E-06 | 5.77E-06 |
| 0.931098 | 0.904586984 | 0.958158198 | DARABCATK12.PWY..D.arabinose.degradation.I | 1.13E-06 | 4.34E-06 |
| 0.9308 | 0.903689163 | 0.958394779 | PWY.7323..superpathway.of.GDP.mannose.derived.O.antigen.building.blocks.biosynthesis | 1.74E-06 | 6.35E-06 |
| 0.930695 | 0.90428483 | 0.957640846 | HEME.BIOSYNTHESIS.II.1..heme.b.biosynthesis.V..aerobic. | 8.94E-07 | 3.56E-06 |
| 0.929855 | 0.902400212 | 0.957862989 | PWY.6859..all.trans.farnesol.biosynthesis | 1.73E-06 | 6.35E-06 |
| 0.929261 | 0.903229953 | 0.955775606 | PWY0.1415..superpathway.of.heme.b.biosynthesis.from.uroporphyrinogen.III | 3.61E-07 | 1.58E-06 |
| 0.928898 | 0.901641833 | 0.956701965 | PWY.5136..fatty.acid..beta..oxidation.II..plant.peroxisome. | 1.06E-06 | 4.09E-06 |
| 0.92869 | 0.901652768 | 0.956316371 | PWY.822..fructan.biosynthesis | 8.25E-07 | 3.32E-06 |
| 0.928393 | 0.898774554 | 0.958632487 | PWY.5692..allantoin.degradation.to.glyoxylate.II | 6.16E-06 | 2.05E-05 |
| 0.928393 | 0.898774554 | 0.958632487 | URDEGR.PWY..superpathway.of.allantoin.degradation.in.plants | 6.16E-06 | 2.05E-05 |
| 0.928339 | 0.901777168 | 0.955430824 | PWY.7187..pyrimidine.deoxyribonucleotides.de.novo.biosynthesis.II | 4.50E-07 | 1.90E-06 |
| 0.927828 | 0.900596772 | 0.955555576 | FUC.RHAMCAT.PWY..superpathway.of.fucose.and.rhamnose.degradation | 7.05E-07 | 2.89E-06 |
| 0.927705 | 0.900948347 | 0.954906208 | COLANSYN.PWY..colanic.acid.building.blocks.biosynthesis | 4.36E-07 | 1.86E-06 |
| 0.926939 | 0.899464593 | 0.955023354 | GLYCOLYSIS.TCA.GLYOX.BYPASS..superpathway.of.glycolysis..pyruvate.dehydrogenase..TCA..and.glyoxylate.bypass | 6.88E-07 | 2.85E-06 |
| 0.926822 | 0.900235849 | 0.953983693 | PWY0.166..superpathway.of.pyrimidine.deoxyribonucleotides.de.novo.biosynthesis..E..coli. | 2.75E-07 | 1.24E-06 |
| 0.926791 | 0.899614432 | 0.954369711 | PWY.1269..CMP.3.deoxy.D.manno.octulosonate.biosynthesis | 4.74E-07 | 1.98E-06 |
| 0.925888 | 0.899697567 | 0.952564549 | PWY.6961..L.ascorbate.degradation.II..bacterial..aerobic. | 1.23E-07 | 5.96E-07 |
| 0.92587 | 0.898788062 | 0.953513855 | PWY.6531..mannitol.cycle | 3.21E-07 | 1.43E-06 |
| 0.925743 | 0.898006598 | 0.953696339 | PWY.621..sucrose.degradation.III..sucrose.invertase. | 4.21E-07 | 1.82E-06 |
| 0.925559 | 0.89715502 | 0.954593136 | KDO.NAGLIPASYN.PWY..superpathway.of..Kdo.2.lipid.A.biosynthesis | 1.01E-06 | 3.95E-06 |
| 0.925353 | 0.897862986 | 0.95329595 | PWY.7663..gondoate.biosynthesis..anaerobic. | 3.40E-07 | 1.50E-06 |
| 0.924989 | 0.897872846 | 0.952628039 | HEME.BIOSYNTHESIS.II..heme.b.biosynthesis.I..aerobic. | 2.39E-07 | 1.10E-06 |
| 0.924652 | 0.898134026 | 0.951716241 | PWY.7883..anhydromuropeptides.recycling.II | 1.15E-07 | 5.62E-07 |
| 0.924609 | 0.89805632 | 0.951674763 | PRPP.PWY..superpathway.of.histidine..purine..and.pyrimidine.biosynthesis | 1.15E-07 | 5.62E-07 |
| 0.924562 | 0.897252833 | 0.952311747 | P441.PWY..superpathway.of.N.acetylneuraminate.degradation | 2.47E-07 | 1.13E-06 |
| 0.92422 | 0.894760736 | 0.954307124 | CARNMET.PWY..L.carnitine.degradation.I | 1.59E-06 | 5.93E-06 |
| 0.923931 | 0.897518722 | 0.950854099 | PWY.5910..superpathway.of.geranylgeranyldiphosphate.biosynthesis.I..via.mevalonate. | 7.65E-08 | 3.84E-07 |
| **0.923564** | **0.896354044** | **0.951180114** | **ARGININE.SYN4.PWY..L.ornithine.biosynthesis.II** | **1.39E-07** | **6.64E-07** |
| 0.923453 | 0.896152859 | 0.951460195 | PWY.6608..guanosine.nucleotides.degradation.III | 1.84E-07 | 8.61E-07 |
| 0.923319 | 0.895961693 | 0.951262006 | ECASYN.PWY..enterobacterial.common.antigen.biosynthesis | 1.75E-07 | 8.26E-07 |
| 0.923156 | 0.89668361 | 0.950136188 | PWY.922..mevalonate.pathway.I..eukaryotes.and.bacteria. | 6.10E-08 | 3.19E-07 |
| 0.922653 | 0.896148361 | 0.949620102 | PWY.5837..2.carboxy.1.4.naphthoquinol.biosynthesis | 5.10E-08 | 2.80E-07 |
| 0.922338 | 0.895447049 | 0.949625336 | HEMESYN2.PWY..heme.b.biosynthesis.II..oxygen.independent. | 7.21E-08 | 3.72E-07 |
| 0.922168 | 0.895765559 | 0.949134064 | PWY.801..homocysteine.and.cysteine.interconversion | 3.99E-08 | 2.40E-07 |
| **0.92169** | **0.894984526** | **0.948858708** | **PWY.5505..L.glutamate.and.L.glutamine.biosynthesis** | **4.51E-08** | **2.63E-07** |
| 0.921515 | 0.894642098 | 0.948910099 | PWY.8178..pentose.phosphate.pathway..non.oxidative.branch..II | 5.12E-08 | 2.80E-07 |
| 0.921501 | 0.89106591 | 0.952561969 | LIPASYN.PWY..phospholipases | 1.53E-06 | 5.77E-06 |
| 0.920988 | 0.89406085 | 0.948204795 | PYRIDOXSYN.PWY..pyridoxal.5..phosphate.biosynthesis.I | 3.98E-08 | 2.40E-07 |
| 0.920553 | 0.894024572 | 0.947538474 | PWY.5861..superpathway.of.demethylmenaquinol.8.biosynthesis.I | 2.34E-08 | 1.49E-07 |
| 0.920534 | 0.893498131 | 0.948122044 | NONOXIPENT.PWY..pentose.phosphate.pathway..non.oxidative.branch..I | 4.50E-08 | 2.63E-07 |
| 0.919918 | 0.892563222 | 0.947763531 | P621.PWY..nylon.6.oligomer.degradation | 4.86E-08 | 2.75E-07 |
| 0.919484 | 0.891910226 | 0.947601238 | PPGPPMET.PWY..ppGpp.metabolism | 5.45E-08 | 2.88E-07 |
| 0.919164 | 0.892036405 | 0.946826912 | PWY66.429..fatty.acid.biosynthesis.initiation..mitochondria. | 2.91E-08 | 1.83E-07 |
| **0.9184** | **0.892061939** | **0.945285453** | **PWY.6293..superpathway.of.L.cysteine.biosynthesis..fungi.** | **8.38E-09** | **5.91E-08** |
| 0.91803 | 0.89067394 | 0.945383938 | PANTOSYN.PWY..superpathway.of.coenzyme.A.biosynthesis.I..bacteria. | 1.52E-08 | 9.83E-08 |
| 0.917973 | 0.891226758 | 0.945231081 | PWY0.301..L.ascorbate.degradation.I..bacterial..anaerobic. | 1.16E-08 | 7.75E-08 |
| 0.91719 | 0.890442058 | 0.94457051 | PANTO.PWY..phosphopantothenate.biosynthesis.I | 9.31E-09 | 6.45E-08 |
| **0.916649** | **0.889769073** | **0.944039675** | **P122.PWY..heterolactic.fermentation** | **8.18E-09** | **5.87E-08** |
| 0.916341 | 0.889098317 | 0.944133008 | PWY.7211..superpathway.of.pyrimidine.deoxyribonucleotides.de.novo.biosynthesis | 1.16E-08 | 7.75E-08 |
| 0.916313 | 0.889019023 | 0.944191907 | PWY0.1298..superpathway.of.pyrimidine.deoxyribonucleosides.degradation | 1.18E-08 | 7.75E-08 |
| 0.915077 | 0.88848554 | 0.942107832 | PWY.5897..superpathway.of.menaquinol.11.biosynthesis | 2.86E-09 | 2.25E-08 |
| 0.915077 | 0.88848554 | 0.942107832 | PWY.5898..superpathway.of.menaquinol.12.biosynthesis | 2.86E-09 | 2.25E-08 |
| 0.915077 | 0.88848554 | 0.942107832 | PWY.5899..superpathway.of.menaquinol.13.biosynthesis | 2.86E-09 | 2.25E-08 |
| 0.914583 | 0.887224479 | 0.942404842 | PWY.5981..CDP.diacylglycerol.biosynthesis.III | 6.47E-09 | 4.73E-08 |
| 0.913222 | 0.886900811 | 0.939969298 | PWY.5845..superpathway.of.menaquinol.9.biosynthesis | 8.99E-10 | 9.27E-09 |
| 0.91313 | 0.885933554 | 0.940550737 | FUCCAT.PWY..fucose.degradation | 2.28E-09 | 1.91E-08 |
| 0.913011 | 0.886174915 | 0.940309479 | GALACTARDEG.PWY..D.galactarate.degradation.I | 1.74E-09 | 1.60E-08 |
| 0.913011 | 0.886174915 | 0.940309479 | GLUCARGALACTSUPER.PWY..superpathway.of.D.glucarate.and.D.galactarate.degradation | 1.74E-09 | 1.60E-08 |
| 0.912434 | 0.886117735 | 0.939164567 | PWY.5862..superpathway.of.demethylmenaquinol.9.biosynthesis | 6.34E-10 | 7.08E-09 |
| 0.911759 | 0.885148396 | 0.938798001 | PWY.5838..superpathway.of.menaquinol.8.biosynthesis.I | 7.41E-10 | 8.05E-09 |
| **0.911719** | **0.884519287** | **0.939335098** | **PWY.7013...S..propane.1.2.diol.degradation** | **1.80E-09** | **1.60E-08** |
| **0.911342** | **0.884020179** | **0.939196395** | **PWY.6936..seleno.amino.acid.biosynthesis..plants.** | **1.86E-09** | **1.63E-08** |
| 0.911124 | 0.883719827 | 0.939019103 | PWY66.409..superpathway.of.purine.nucleotide.salvage | 1.91E-09 | 1.64E-08 |
| **0.91071** | **0.883678378** | **0.937975607** | **PWY.5154..L.arginine.biosynthesis.III..via.N.acetyl.L.citrulline.** | **8.90E-10** | **9.27E-09** |
| 0.910609 | 0.883261261 | 0.938463944 | GLCMANNANAUT.PWY..superpathway.of.N.acetylglucosamine..N.acetylmannosamine.and.N.acetylneuraminate.degradation | 1.29E-09 | 1.30E-08 |
| 0.910281 | 0.879721238 | 0.941397236 | PWY0.1533..methylphosphonate.degradation.I | 5.23E-08 | 2.80E-07 |
| 0.910281 | 0.879721238 | 0.941397236 | PWY.7807..glyphosate.degradation.III | 5.23E-08 | 2.80E-07 |
| 0.907983 | 0.879973376 | 0.935922901 | PWY.7282..4.amino.2.methyl.5.diphosphomethylpyrimidine.biosynthesis.II | 2.68E-10 | 3.53E-09 |
| 0.907689 | 0.880646805 | 0.934786659 | PWY0.1586..peptidoglycan.maturation..meso.diaminopimelate.containing. | 2.51E-10 | 3.47E-09 |
| 0.906107 | 0.879747414 | 0.932952366 | P105.PWY..TCA.cycle.IV..2.oxoglutarate.decarboxylase. | 4.53E-11 | 7.92E-10 |
| 0.905644 | 0.877999542 | 0.933555319 | PWY0.1261..anhydromuropeptides.recycling.I | 2.72E-10 | 3.53E-09 |
| 0.905358 | 0.87892049 | 0.932135927 | PWY.5840..superpathway.of.menaquinol.7.biosynthesis | 3.27E-11 | 6.57E-10 |
| **0.905058** | **0.878281633** | **0.932353223** | **P4.PWY..superpathway.of.L.lysine..L.threonine.and.L.methionine.biosynthesis.I** | **5.81E-11** | **8.98E-10** |
| 0.905058 | 0.878281633 | 0.932353223 | PWY0.781..aspartate.superpathway | 5.81E-11 | 8.98E-10 |
| 0.904974 | 0.878247275 | 0.932230716 | P164.PWY..purine.nucleobases.degradation.I..anaerobic. | 5.32E-11 | 8.92E-10 |
| 0.903779 | 0.87690119 | 0.931006475 | PWY.6305..superpathway.of.putrescine.biosynthesis | 3.74E-11 | 7.15E-10 |
| 0.903673 | 0.875409349 | 0.932439558 | PWY.7269..mitochondrial.NADPH.production..yeast. | 3.06E-10 | 3.84E-09 |
| 0.903406 | 0.87602823 | 0.930917008 | POLYISOPRENSYN.PWY..polyisoprenoid.biosynthesis..E..coli. | 4.52E-11 | 7.92E-10 |
| 0.902091 | 0.87514117 | 0.929535579 | FOLSYN.PWY..superpathway.of.tetrahydrofolate.biosynthesis.and.salvage | 2.06E-11 | 4.59E-10 |
| 0.902091 | 0.87514117 | 0.929535579 | PWY.6612..superpathway.of.tetrahydrofolate.biosynthesis | 2.06E-11 | 4.59E-10 |
| 0.901845 | 0.87483732 | 0.929251636 | PWY.7199..pyrimidine.deoxyribonucleosides.salvage | 1.57E-11 | 3.95E-10 |
| 0.901641 | 0.874726759 | 0.929007987 | PWY0.1477..ethanolamine.utilization | 1.57E-11 | 3.95E-10 |
| 0.89785 | 0.870330175 | 0.924961653 | PWY.5659..GDP.mannose.biosynthesis | 1.88E-12 | 6.31E-11 |
| 0.89681 | 0.866001727 | 0.927988464 | PROTOCATECHUATE.ORTHO.CLEAVAGE.PWY..protocatechuate.degradation.II..ortho.cleavage.pathway. | 6.28E-10 | 7.08E-09 |
| 0.895506 | 0.868633711 | 0.92279343 | GLUCARDEG.PWY..D.glucarate.degradation.I | 8.25E-13 | 3.01E-11 |
| 0.895474 | 0.867895239 | 0.923472712 | PWY0.1297..superpathway.of.purine.deoxyribonucleosides.degradation | 3.41E-12 | 1.06E-10 |
| 0.882976 | 0.855955079 | 0.910385067 | MET.SAM.PWY..superpathway.of.S.adenosyl.L.methionine.biosynthesis | 2.42E-15 | 1.08E-13 |
| **0.882976** | **0.855955079** | **0.910385067** | **PWY.5347..superpathway.of.L.methionine.biosynthesis..transsulfuration.** | **2.42E-15** | **1.08E-13** |
| **0.878828** | **0.851941569** | **0.906067325** | **HOMOSER.METSYN.PWY..L.methionine.biosynthesis.I** | **1.96E-16** | **1.31E-14** |
| **0.87844** | **0.851494639** | **0.905740391** | **METSYN.PWY..superpathway.of.L.homoserine.and.L.methionine.biosynthesis** | **1.86E-16** | **1.31E-14** |
| 0.878269 | 0.850799394 | 0.905992406 | PHOSLIPSYN.PWY..superpathway.of.phospholipid.biosynthesis.I..bacteria. | 4.62E-16 | 2.65E-14 |
| **0.876771** | **0.849668076** | **0.904151444** | **P461.PWY..hexitol.fermentation.to.lactate..formate..ethanol.and.acetate** | **8.14E-17** | **1.31E-14** |
| 0.876661 | 0.849235545 | 0.904377599 | PWY4FS.7..phosphatidylglycerol.biosynthesis.I..plastidic. | 1.90E-16 | 1.31E-14 |
| 0.876661 | 0.849235545 | 0.904377599 | PWY4FS.8..phosphatidylglycerol.biosynthesis.II..non.plastidic. | 1.90E-16 | 1.31E-14 |
| 0.873022 | 0.846220062 | 0.900056733 | RHAMCAT.PWY..L.rhamnose.degradation.I | 4.49E-18 | 1.81E-15 |

**Supplementary Table 3.** The differences in abundance of microbial functional capacity for all mapped metagenomic pathways at the whole community level in residents of long-term aged care facilities with increasingly severe CI, at statistical significance. The magnitude of the difference is presented as highest to lowest odds ratio with 95% confidence intervals following a multivariate adjustment for time since cognitive impairment assessment, age, sex, antibiotic use, proton pump inhibitor use, opioid use, laxative use, recorded medical history, meal texture, and liquid texture. Pathways of interest identified in bold.

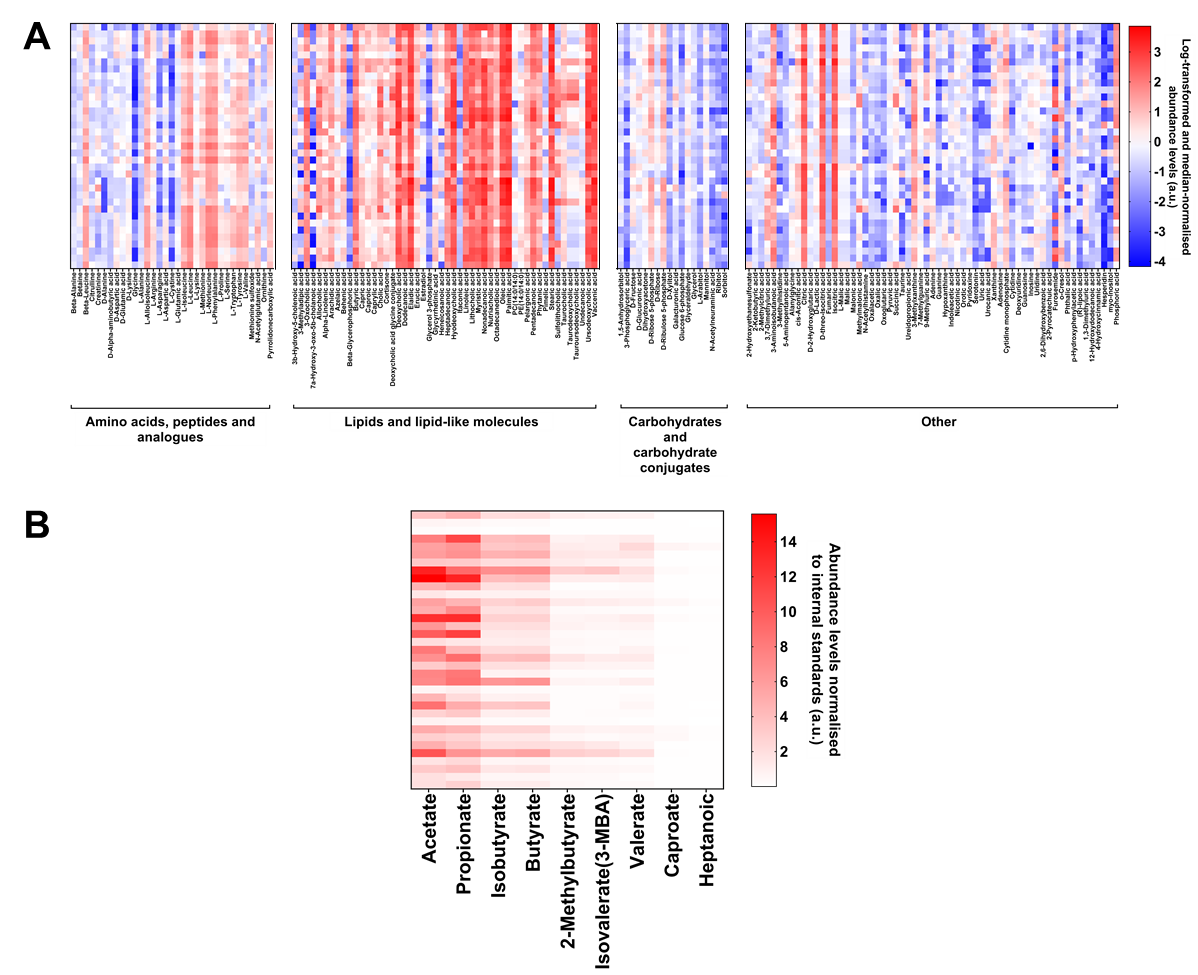

**Supplementary Figure 6. Metabolite profiles of cognitively impaired residents of long-term aged care facilities.** Heatmaps illustrating the detected levels of **A**) 165 polar metabolites including amino acids, peptides and analogues, lipids and lipid-like molecules, carbohydrates and carbohydrate conjugates, and other metabolites (organic acids, organoheterocyclic compounds, nucleosides, nucleotides and analogues, benzenoids, medium-chain fatty acids, phenylpropanoids and polyketides, organic oxygen compounds, and inorganic compounds), and **B**) nine short-chain fatty acids.

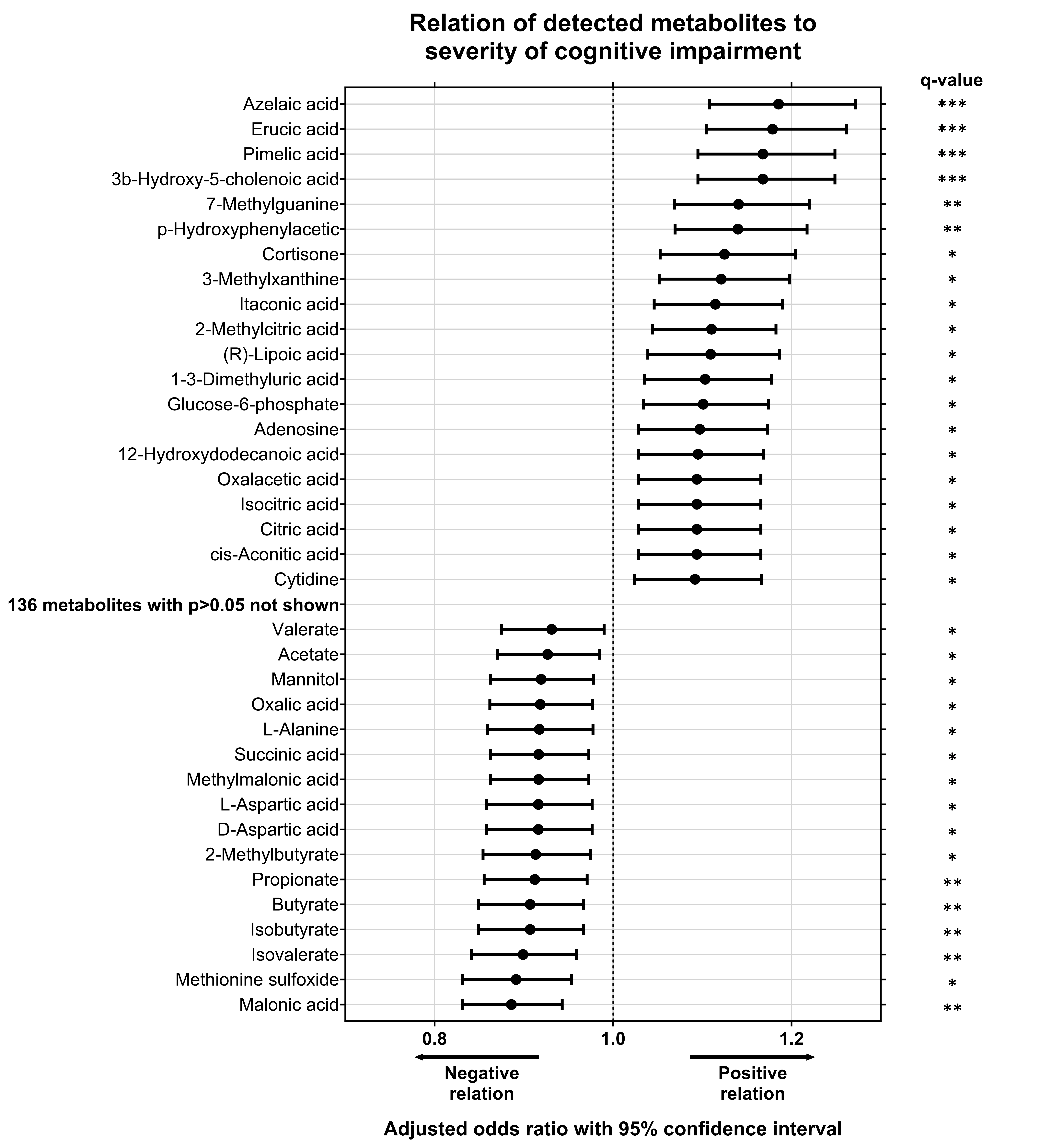

**Supplementary Figure 7. Metabolite differences in the gut microbiome of residents of long-term aged care by cognitive impairment.** All metabolites detected with significant differences by CI severity include short-chain fatty acids and polar metabolites. Odds ratio and 95% confidence interval of effect of cognitive impairment severity on detected metabolite abundance levels. Performed by multivariate analysis, adjusting for time since cognitive impairment assessment, age, sex, antibiotic use, proton pump inhibitor use, opioid use, laxative use, recorded medical history, meal texture, and liquid texture. n=12 mild; n=11 moderate; n=12 severe. *q<0.05; **q<0.01; ***q<0.001 for adjusted p-values following FDR correction.

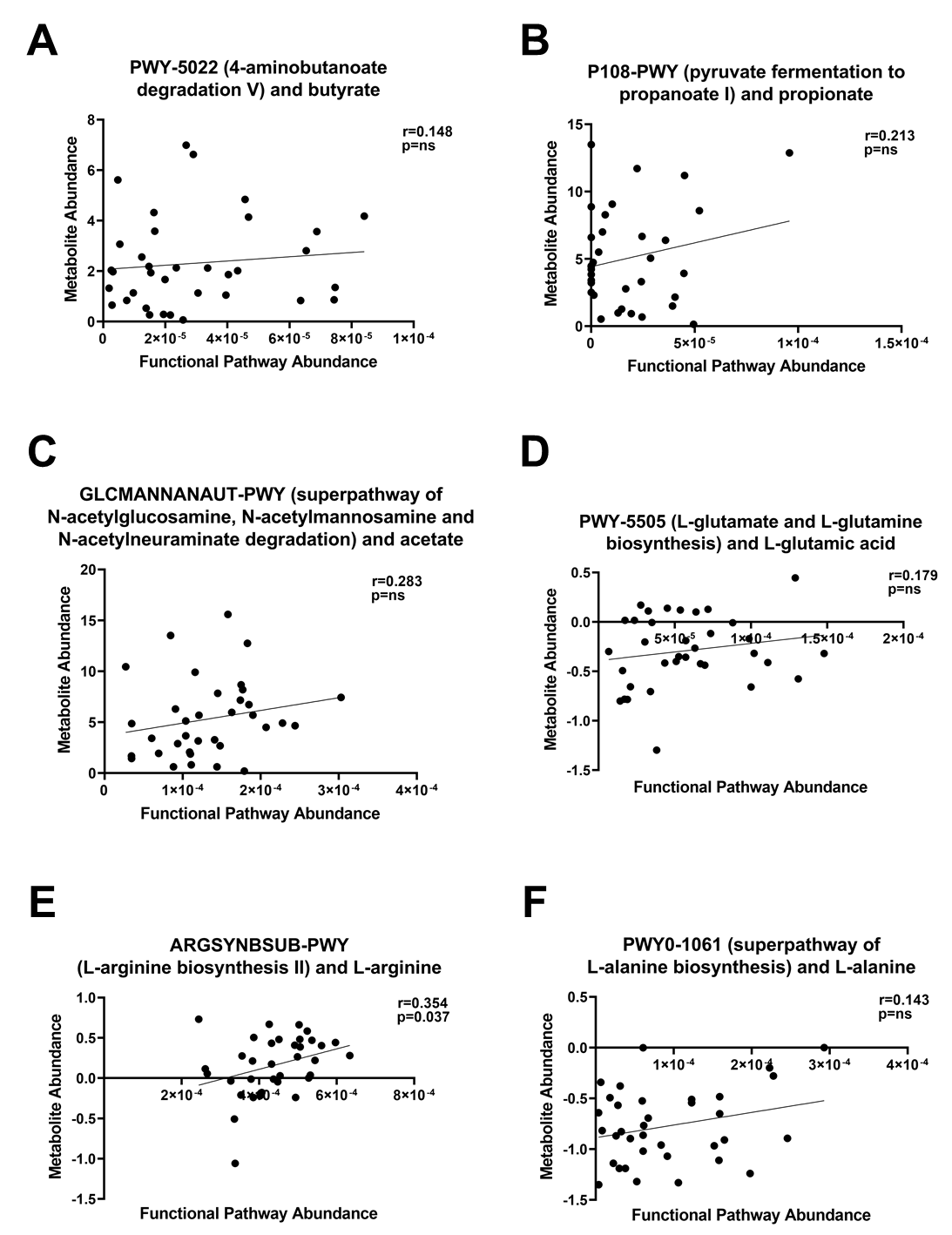

**Supplementary Figure 8. Microbial functional capacity correlates with detected metabolite levels in the gut microbiome of residents in long-term aged care with cognitive impairment.** Correlations of the relative abundance of microbial functional capacity for mapped metagenomic pathways at the whole community level with direct metabolites detected in stool samples of the CI cohort, determined with Spearman’s correlation. n=12 mild; n=11 moderate; n=12 severe.
